# Effects of Screening for Colorectal Cancer: Development, Documentation and Validation of a Multistate Markov Model

**DOI:** 10.1101/2020.04.17.20069484

**Authors:** Thomas Heisser, Michael Hoffmeister, Hermann Brenner

**Author notes:** Correspondence: Thomas Heisser, Division of Clinical Epidemiology and Aging Research, German Cancer Research Center (DKFZ), Im Neuenheimer Feld 581, 69120 Heidelberg, Germany. Categories: Innovative Tools and Methods; Cancer Epidemiology; Cancer Therapy and Prevention.

## Abstract

Simulation models are a powerful tool to overcome gaps of evidence needed to inform medical decision making. Here, we present development and application of a multistate Markov model to simulate effects of colorectal cancer (CRC) screening, along with a thorough assessment of the model’s ability to reproduce real-life outcomes. Firstly, we provide a comprehensive documentation of the model development, structure and assumptions. Secondly, to assess the model’s external validity, we compared model-derived cumulative incidence and prevalences of colorectal neoplasms to (1) results from KolosSal, a study in German screening colonoscopy participants, (2) registry-based estimates of CRC incidence in Germany, and (3) outcome patterns of randomized sigmoidoscopy screening studies. We found that (1) more than 90% of observed prevalences in the KolosSal study were within the 95% confidence intervals of the model-predicted neoplasm prevalences; (2) the 15-year cumulative CRC incidences estimated by simulations for the German population deviated by 0.0-0.2 percent units in men and 0.0-0.3 percent units in women when compared to corresponding registry-derived estimates; and (3) the time course of cumulative CRC incidence and mortality in the modelled intervention group and control group closely resembles the time course reported from sigmoidoscopy screening trials. Summarized, our model adequately predicted colorectal neoplasm prevalences and incidences in a German population for up to 25 years, with estimated patterns of the effect of screening colonoscopy resembling those seen in registry data and real-world studies. This suggests that the model may represent a valid tool to assess the comparative effectiveness of CRC screening strategies.

**Novelty and Impact:** The best long-term strategy to screen for colorectal cancer is unknown. To overcome gaps of evidence in this context, we developed a simulation model and assessed its external validity. The model predicted the natural history of colorectal cancer for up to 25 years and was able to reproduce effects of screening seen in real-world data. Our model may be used as a transparent, valid tool to inform medical decision making on colorectal cancer screening strategies.

## Introduction

Colorectal cancer (CRC) is one of few cancers for which effective screening options are established. While population-based CRC screening programs are already in place or being rolled out in numerous countries worldwide,^1^ health authorities remain concerned with identifying screening strategies which optimally balance health outcomes against resource use and test burden. This, however, is a challenging endeavor, as screening modalities vary largely in diagnostic performance, uptake, test burden, and costs. Combined, these factors allow for a multitude of possible screening strategies, only some of which have by now been assessed in randomized or observational studies, with inevitably limited scope of assessed strategies and follow-up periods.^2–11^

Simulation models are a powerful tool to overcome these evidence gaps required to inform medical decision-making. However, as models are at least as imperfect as the real-world data used for their development, a model’s success depends on the levels of trust decision-makers will have in its results.^12^ To enhance a model’s credibility, firstly, transparency regarding the model’s structure, underlying parameters and assumptions is crucial. Secondly, a model should be subjected to a validation process by measuring modelled outcomes against observed real-world data the model ought to reproduce. Such external validation is regarded a particular robust form of validation and thus an indispensable step in model development.^12^

In several previous analysis, we step-by-step developed a multistate Markov model to simulate effects of CRC screening in a hypothetical population.^13–16^ The objective of the present study was, firstly, to provide a thorough and transparent documentation of the model development, structure and assumptions, enabling interested readers to genuinely understand the model’s functionality and potentially even to conduct their own analysis. Secondly, we sought to comprehensively address the model’s ability to reproduce real-world outcomes by a three-way-validation approach: we compared model-derived outcomes to (1) results from a German colonoscopy screening study (KolosSal study), (2) estimates based on German registry data, and (3) outcome patterns reported from randomized controlled trials (RCTs) on the effects of screening sigmoidoscopy.

## Material and Methods

### Model Documentation

A documentation on the model’s structure and data sources used for its development is provided in **Supplementary Appendix 1a**, including overviews on all model parameters (**Supplementary Tables 1-3**). Briefly, our multistate Markov model simulates the natural history of CRC in a hypothetical population for a predefined number of years. Screening can interfere with the natural history of CRC (**Figure 1**).

**Table 1.**
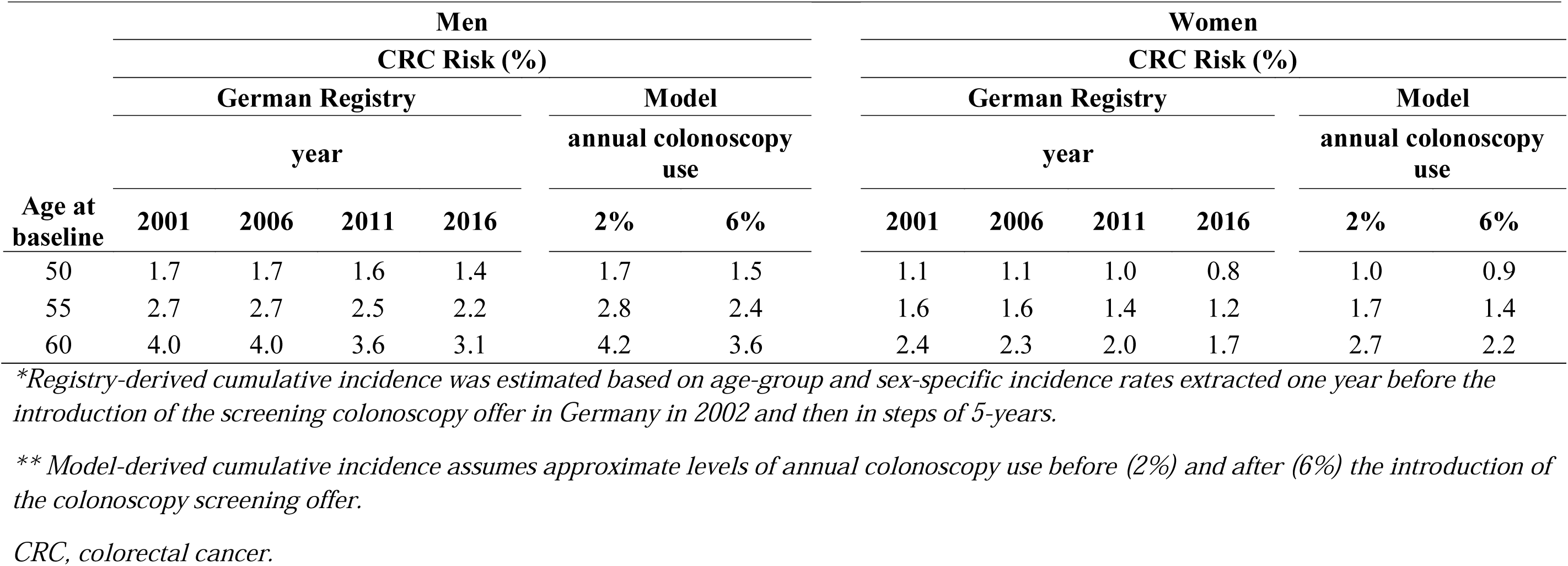
Registry*-vs. model**-derived cumulative incidence of colorectal cancer after 15 years from baseline for ages 50-64, 55-69, and 60-74

| Age at<br>baseline | Men |  |  |  |  |  | Women |  |  |  |  |  |
| --- | --- | --- | --- | --- | --- | --- | --- | --- | --- | --- | --- | --- |
|  | CRC Risk (%) |  |  |  |  |  | CRC Risk (%) |  |  |  |  |  |
|  | German Registry |  |  |  | Model |  | German Registry |  |  |  | Model |  |
|  | year |  |  |  | annual colonoscopy<br>use |  | year |  |  |  | annual colonoscopy<br>use |  |
|  | 2001 | 2006 | 2011 | 2016 | 2% | 6% | 2001 | 2006 | 2011 | 2016 | 2% | 6% |
| 50 | 1.7 | 1.7 | 1.6 | 1.4 | 1.7 | 1.5 | 1.1 | 1.1 | 1.0 | 0.8 | 1.0 | 0.9 |
| 55 | 2.7 | 2.7 | 2.5 | 2.2 | 2.8 | 2.4 | 1.6 | 1.6 | 1.4 | 1.2 | 1.7 | 1.4 |
| 60 | 4.0 | 4.0 | 3.6 | 3.1 | 4.2 | 3.6 | 2.4 | 2.3 | 2.0 | 1.7 | 2.7 | 2.2 |

\*Registry-derived cumulative incidence was estimated based on age-group and sex-specific incidence rates extracted one year before the introduction of the screening colonoscopy offer in Germany in 2002 and then in steps of 5-years.
\*\* Model-derived cumulative incidence assumes approximate levels of annual colonoscopy use before (2%) and after (6%) the introduction of the colonoscopy screening offer.
CRC, colorectal cancer.

**Figure 1.**
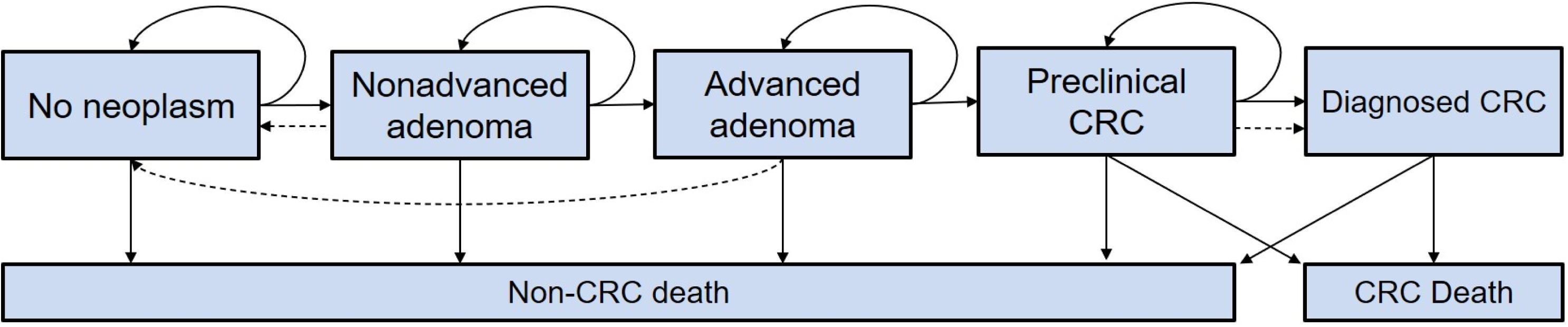
Schematic illustration of the Markov Model. *Solid lines represent the progression of colorectal disease through the adenoma-carcinoma sequence in the absence of screening; dashed lines show the movement between states because of the detection and removal of adenomas and the detection of asymptomatic CRC at screening*. *CRC, colorectal cancer*.

The analyses presented in this study are based on simulations for a hypothetical previously unscreened German population, in line with the used input parameters derived from Germany-centred data sources. The model can principally be used for simulating any population, provided updated or appropriately adjusted input parameters. The model was developed in the open-source statistical software R (version 4.0.2). The model source code is available in **Supplementary Appendix 1b**.

### Model Validation

Details on data sources as well as a comprehensive outline of statistical methods used for the three-fold validation process can be found in **Supplementary Appendix 1c**. In brief, we firstly used our model to simulate prevalences of adenomas and cancers detected at screening colonoscopy by sex and age group (ages 55-75) and compared these to observed prevalences in the KolosSal study, a statewide cohort study in the German state of Saarland (validation part 1). The objective was to assess the agreement between modelled and observed prevalences of any neoplasm (ANN, any adenomas and cancers) and of any advanced neoplasm (ADN, advanced adenomas and cancer). We therefore calculated the prevalence ratios (PRR) by dividing the modelled prevalence estimates by the prevalences observed in KolosSal, both age-specific and as meta-analyses combining all sex-specific PRRs. Statistical equivalence between the modelled and observed prevalences was then tested by applying two one-sided t-tests (TOST)^17^ to each of the sex-specific meta-analyses estimates at a 20% equivalence margin.

Secondly, we simulated populations with levels of annual colonoscopy use representative for the German general population and compared the resulting cumulative incidences to those derived from German registry data before and after the introduction of a population-wide colonoscopy screening offer in the year 2002 (validation part 2, **Supplementary Table 4**). The objective was to assess the agreement between modelled and observed 15-year cumulative CRC incidences for different ages (50-64, 55-59 and 60-74) over time. For the modelled population, we assumed 2% and 6% annual colonoscopy use before and after the introduction of the colonoscopy screening offer, respectively. These utilization levels were chosen as the estimated use of colonoscopy for any reason within 10 years in Germany was 24.3% and 63.9% for the periods 2000-2002 and 2008-2010, respectively, reflecting the increase triggered by the introduction of the colonoscopy screening offer in 2002.^18^ Model starting prevalences were discounted by 30% in order to increase comparability to the German general population given that, already in the period 2000-2002, approximately 66% of 55-59 year-olds had either fecal testing within two years or colonoscopy within ten years.^18^

Thirdly, we simulated a hypothetical colonoscopy screening RCT by modelling exemplary populations with and without baseline colonoscopy screening and assessed the CRC incidence and mortality over time (validation part 3). The objective was to assess the concordance of the resulting patterns when compared to the results of the screening sigmoidoscopy RCTs (the Italian Screening for COlon REctum (SCORE) trial,^2^ the Norwegian Colorectal Cancer Prevention (NORCCAP) trial,^6,19^ the UK Flexible Sigmoidoscopy Screening (UKFSS) trial,^5^ and the US Prostate, Lung, Colorectal and Ovarian (PLCO) screening trial^7^).

### Sensitivity Analyses

To account for uncertainty related to assumptions on pre-screening in the modelled populations, we conducted a probabilistic sensitivity analysis (PSA) for the comparison of model-derived to registry-derived incidences. The PSA was implemented by replacing the point estimates of transition rates and discounted start prevalences by random draws in 1000 model runs. Then, 95% prediction intervals were derived by assessing the 2.5% and 97.5% quantiles of resulting estimates for model-derived cumulative incidences after 15 years. Notably, as this study’s objective was not to make future predictions (but to assess the external validity of the model’s structure and core parameters), we did not conduct sensitivity analysis for validation part 1 (where we had full access to the data of the KolosSal study) and 3 (where the validation objective was to compare graphical patterns).

## Results

### Part 1: Equivalence to KolosSal Study Prevalences

Overall, 11,912 participants (5,916 men and 5,996 women) of the KolosSal study were included in the analysis (**Supplementary Table 5)**. The large majority (overall, 75 out of 80) of observed ANN and ADN prevalences in the KolosSal study were within the 95% confidence intervals (CIs) of the model-predicted prevalences (**Figure 2**). **Figure 3** shows the corresponding PRR per age-group. Estimates from the multilevel meta-analysis yielded PPR of 1.03 (90% CIs, 0.99 – 1.07) and 1.05 (0.98 – 1.12) for ANN and of 0.96 (0.86 – 1.05) and 1.06 (0.96 – 1.16) for ADN for men and women, respectively. When tested for statistical equivalence at a 20% margin, all TOSTs yielded p-values <0.05 (**Supplementary Figure 1)**.

**Figure 2.**
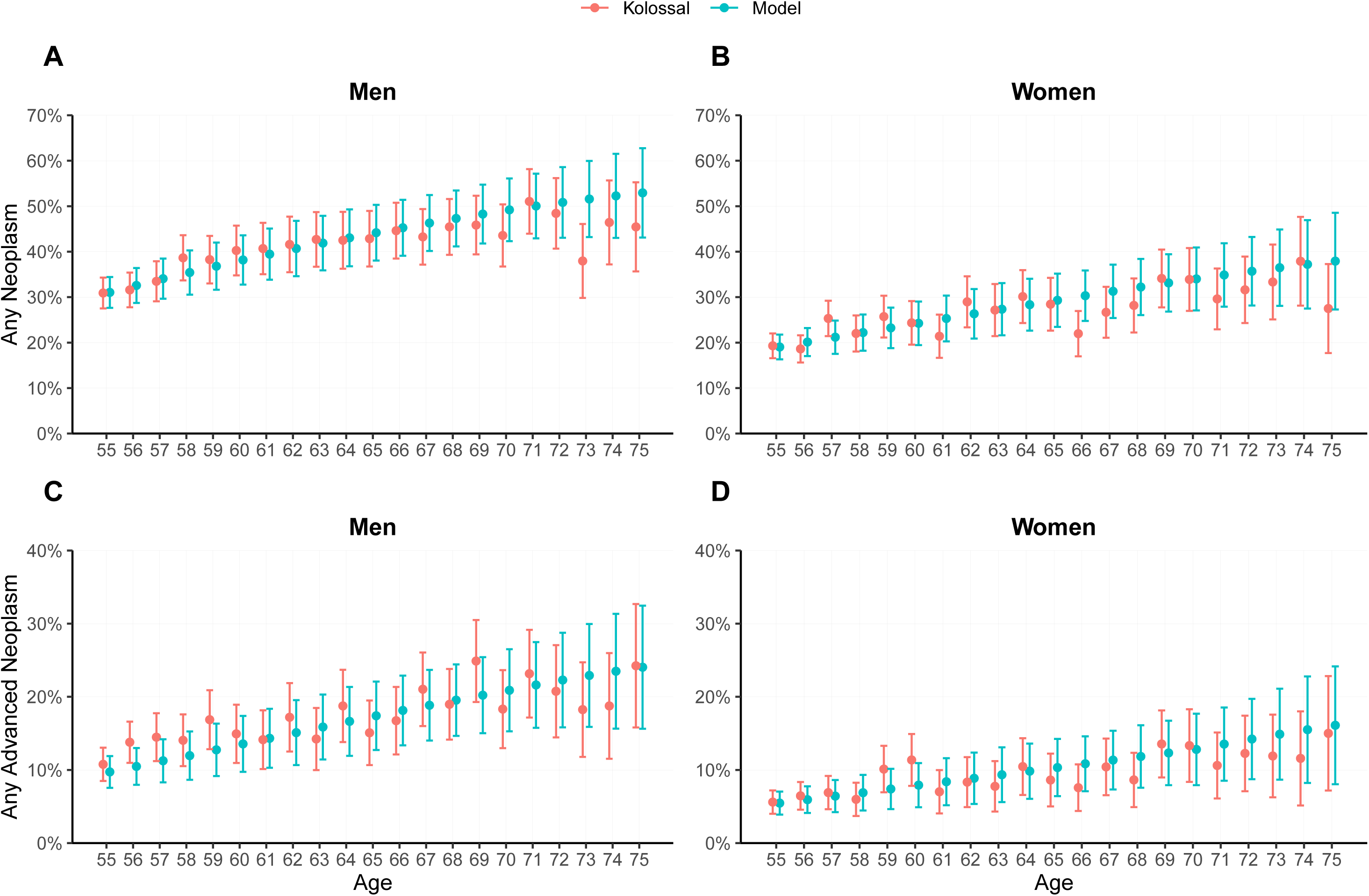
Prevalences and 95% confidence bands for any neoplasms (any adenoma or cancer, panels A and B) and advanced neoplasms (advanced adenomas or cancer, panels C and D) for ages 55-75 in the KolosSal study and as estimated by the simulation model. Left side: men. Right side: women.

**Figure 3.**
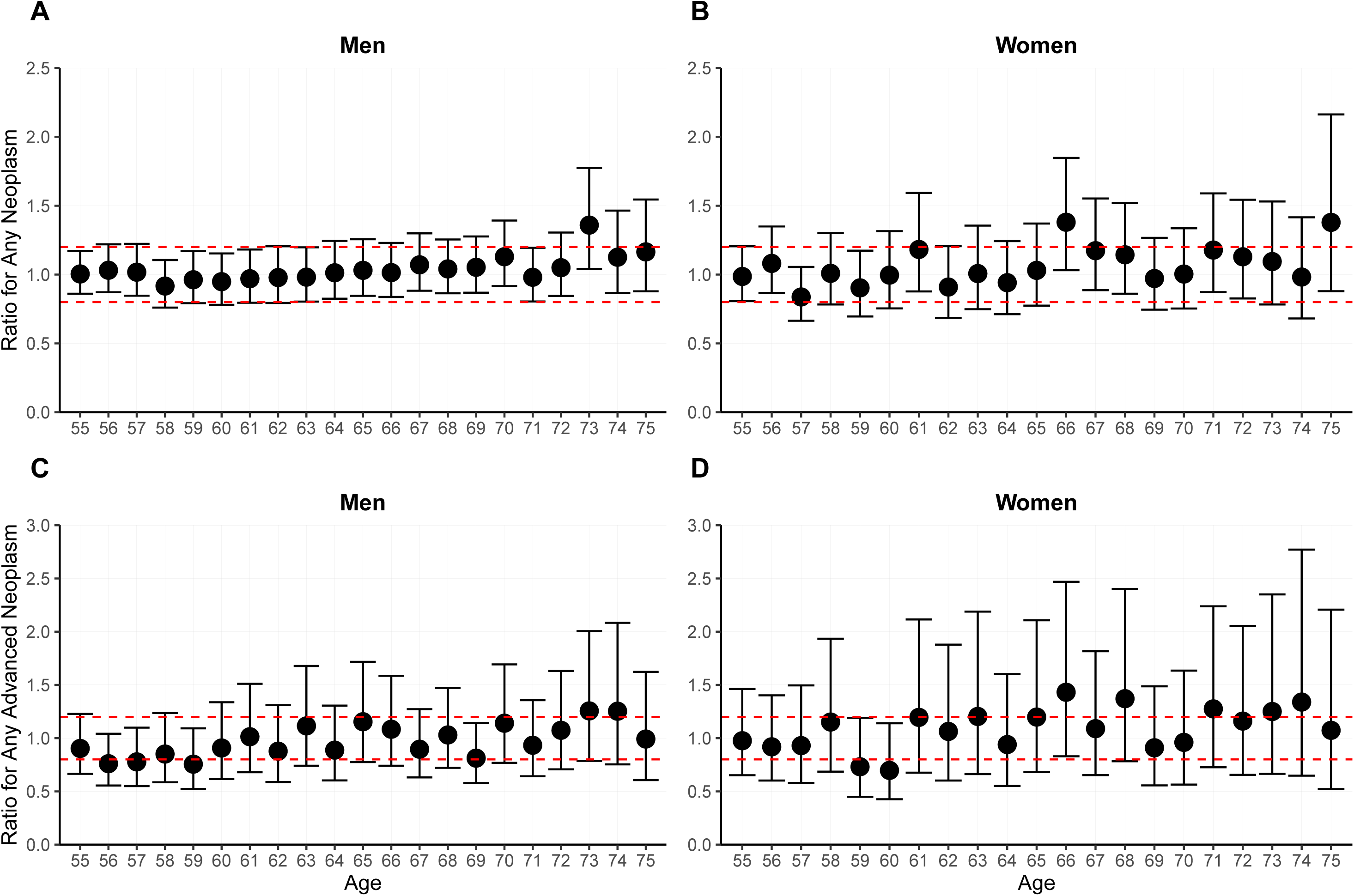
Prevalence ratios and corresponding confidence bands of the estimated prevalences by the simulation model vs the prevalences in KolosSal for ages 55-75 (model estimates divided by prevalences in KolosSal). A, B: any neoplasms. C, D, Any advanced neoplasms. Left side: men. Right side: women. Dotted red lines indicate the margin of testing for equivalence (0.2).

### Part 2: Comparison to Registry-Derived Cumulative Incidences

**Table 1** shows the 15-year registry-vs model-derived cumulative CRC incidences. Overall, the model-derived cumulative incidence with 2% annual colonoscopy use compared best against the registry-derived cumulative incidence based on the year 2001 (deviation of 0.0-0.3 percent units), while the model with 6% annual colonoscopy use was closest to the registry-derived estimate for the year 2011 (deviation of 0.0-0.2 percent units). Deviations between registry- and model-based cumulative incidences tended to increase for older age groups.

### Part 3: Comparison to RCT-Reported CRC Incidence and Mortality Patterns

**Table 2** shows the 15-year risk of developing and dying of CRC for control vs intervention groups, and the effect of screening colonoscopy at baseline. **Supplementary Figure 2** shows the numbers needed to screen to prevent one colorectal cancer event as estimated by the model.

**Table 2.**
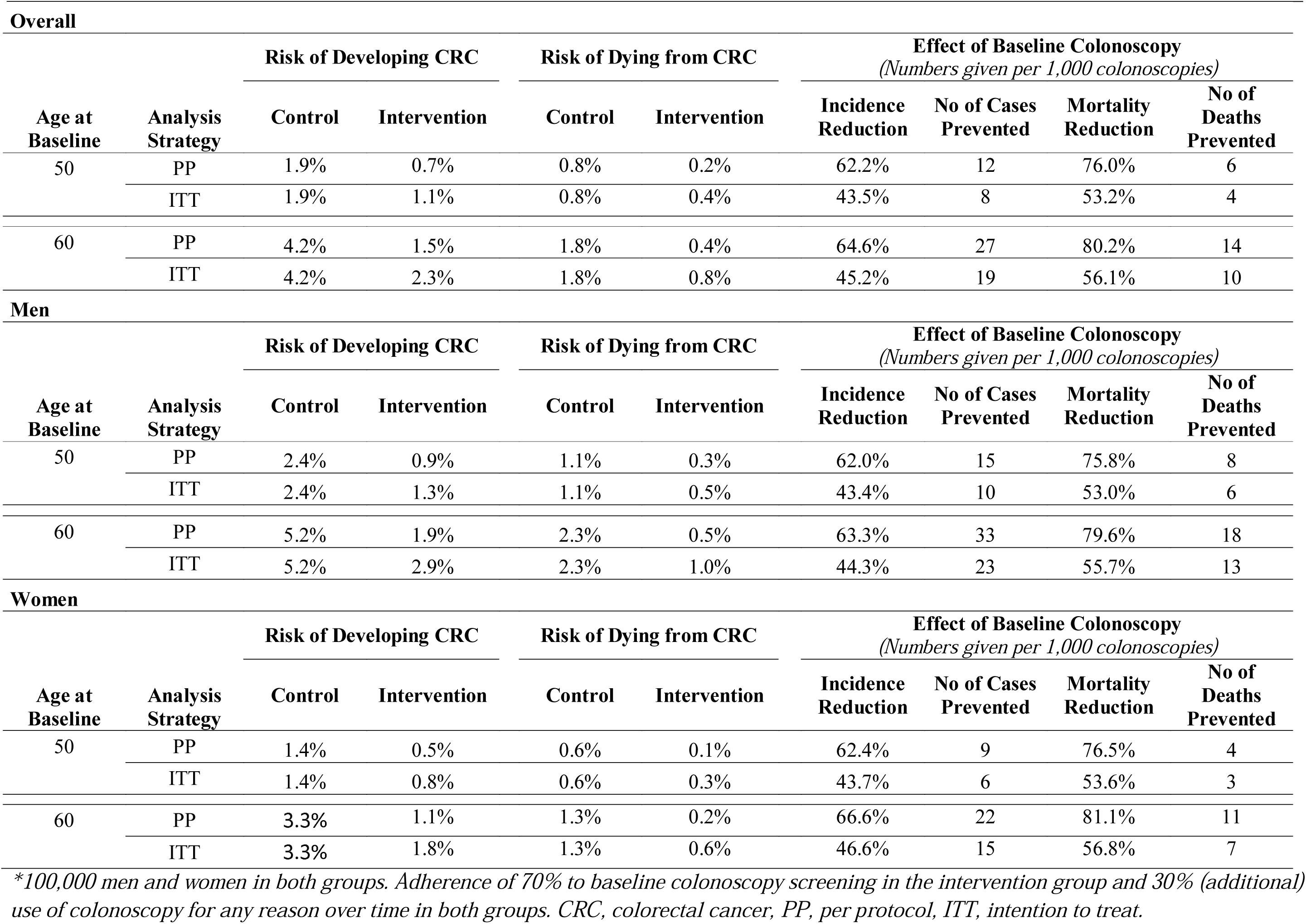
Risk of developing and dying from CRC, and the effect of screening colonoscopy at baseline for modelled control vs intervention groups* with different starting ages after 15 years of follow-up. Shown are results analysed according to the per protocol (PP, only those adherent) and intention to treat (ITT, all randomized) principle

| **Overall** |  |  |  |  |  |  |  |  |  |
| --- | --- | --- | --- | --- | --- | --- | --- | --- | --- |
| Age at Baseline | Analysis Strategy | Risk of Developing CRC |  | Risk of Dying from CRC |  | Effect of Baseline Colonoscopy<br>(Numbers given per 1,000 colonoscopies) |  |  |  |
|  |  | Control | Intervention | Control | Intervention | Incidence Reduction | No of Cases Prevented | Mortality Reduction | No of Deaths Prevented |
| 50 | PP | 1.9% | 0.7% | 0.8% | 0.2% | 62.2% | 12 | 76.0% | 6 |
|  | ITT | 1.9% | 1.1% | 0.8% | 0.4% | 43.5% | 8 | 53.2% | 4 |
| 60 | PP | 4.2% | 1.5% | 1.8% | 0.4% | 64.6% | 27 | 80.2% | 14 |
|  | ITT | 4.2% | 2.3% | 1.8% | 0.8% | 45.2% | 19 | 56.1% | 10 |
| **Men** |  |  |  |  |  |  |  |  |  |
| Age at Baseline | Analysis Strategy | Risk of Developing CRC |  | Risk of Dying from CRC |  | Effect of Baseline Colonoscopy<br>(Numbers given per 1,000 colonoscopies) |  |  |  |
|  |  | Control | Intervention | Control | Intervention | Incidence Reduction | No of Cases Prevented | Mortality Reduction | No of Deaths Prevented |
| 50 | PP | 2.4% | 0.9% | 1.1% | 0.3% | 62.0% | 15 | 75.8% | 8 |
|  | ITT | 2.4% | 1.3% | 1.1% | 0.5% | 43.4% | 10 | 53.0% | 6 |
| 60 | PP | 5.2% | 1.9% | 2.3% | 0.5% | 63.3% | 33 | 79.6% | 18 |
|  | ITT | 5.2% | 2.9% | 2.3% | 1.0% | 44.3% | 23 | 55.7% | 13 |
| **Women** |  |  |  |  |  |  |  |  |  |
| Age at Baseline | Analysis Strategy | Risk of Developing CRC |  | Risk of Dying from CRC |  | Effect of Baseline Colonoscopy<br>(Numbers given per 1,000 colonoscopies) |  |  |  |
|  |  | Control | Intervention | Control | Intervention | Incidence Reduction | No of Cases Prevented | Mortality Reduction | No of Deaths Prevented |
| 50 | PP | 1.4% | 0.5% | 0.6% | 0.1% | 62.4% | 9 | 76.5% | 4 |
|  | ITT | 1.4% | 0.8% | 0.6% | 0.3% | 43.7% | 6 | 53.6% | 3 |
| 60 | PP | 3.3% | 1.1% | 1.3% | 0.2% | 66.6% | 22 | 81.1% | 11 |
|  | ITT | 3.3% | 1.8% | 1.3% | 0.6% | 46.6% | 15 | 56.8% | 7 |

\*100,000 men and women in both groups. Adherence of 70% to baseline colonoscopy screening in the intervention group and 30% (additional) use of colonoscopy for any reason over time in both groups. CRC, colorectal cancer, PP, per protocol, ITT, intention to treat.

Analogous to the overall patterns seen in the SCORE, NORCCAP, UKFSS and PLCO trials on sigmoidoscopy **(Supplementary Figure 3)**, the cumulative incidence in the colonoscopy screening group increased markedly in the year following the screening, followed by modest further increase until the end of follow-up (**Figure 4, Supplementary Figure 4**). In the control group, the cumulative incidence followed a steady growth trajectory. Both curves crossed approximately after 4-5 years, with the gap between curves widening up with longer follow-up duration, a pattern also observed in SCORE, UKFSS and PLCO (in NORCCAP, curves cross approximately after 6 years). While the cumulative mortality in both groups developed almost in parallel in the first years, the curves diverged more markedly with increasing follow-up duration, seemingly exceeding the trends seen in the sigmoidoscopy RCTs. In the model, men and women benefitted at a similar magnitude from the effects of colonoscopy screening (**Supplementary Figures 5-8)**.

**Figure 4.** Per-protocol analysis of cumulative incidence (A, B) and mortality (C, D) in simulated cohorts of each 100,000 men and women in an unscreened control vs an intervention group with baseline colonoscopy screening. Shown are different starting ages at start of the simulation. All cohorts had 16 years of follow-up.

### Sensitivity Analyses

All 95% prediction intervals for modelled cumulative incidences covered the registry-derived incidences for the corresponding years, i.e. 2% and 6% annual colonoscopy use for the years 2001 and 2011, respectively (**Supplementary Table 6**). While accounting for the uncertainty related to the extent of previous screening resulted in relatively large intervals (absolute range, 0.5 – 0.9 % in men, 0.3 – 0.5% in women), model-estimated reductions in cumulative incidences (i.e. when comparing 6% and 2% utilization models) appeared stable over time.

## Discussion

In this study, we firstly provided a comprehensive and transparent documentation of the development, structure and assumption of our multistate Markov model on the effects of CRC screening. Secondly, we closely scrutinized the model’s external validity. Overall, we found (1) a high level of agreement between model-derived and observed prevalences of colorectal neoplasms in a large screening colonoscopy study from Germany; (2) closely matching model- and registry-derived cumulative CRC incidences, absolute and in relative changes over time; and (3) similar trends in outcomes for a hypothetical colonoscopy screening RCT as compared to real-life sigmoidoscopy screening RCTs. Combined, our results suggest that the model may be able to adequately reproduce both the natural history of CRC as well as the effects of colonoscopy screening.

### Validity of the Simulated Natural History

Parts 1 and 2 of the validation, based on the KolosSal study and the German cancer registry, illustrated the model’s ability to predict colorectal neoplasm prevalences and cumulative cancer incidences for up to 25 years. In particular, >90% of observed prevalences in the KolosSal study were within the 95% CIs of the model-predicted prevalences. Analysis of age-specific prevalence ratios suggested that the model might partly overestimate prevalences of advanced neoplasms in older men and women. However, the sample size in KolosSal for age groups >70 years, where model predictions tended to yield higher than observed prevalences, was small (N < 200), increasing the associated uncertainty. In the meta-analysis, explorative testing with two one-sided t-tests suggested statistical equivalence at a 20% equivalence margin for all outcomes.

In addition, a model with 2% annual colonoscopy use closely approximated the registry-derived cumulative CRC incidence for the year 2001, i.e. before colonoscopy screening was introduced. Model-derived incidences for 55-69 and 60-74-year-olds tended to slightly overestimate the registry-derived incidences. A likely explanation is that we assumed constant annual utilization patterns over time for the sake of simplicity, which does not consider that, in the period 2000-2002, older age groups were more likely to use colonoscopy than younger ones.^18^ Interestingly, the cumulative incidence for a model with 6% annual colonoscopy use matched best with the registry-derived estimate for the year 2011. This seems plausible given that the 6% utilization rate was derived for the period 2008-2010. More recent evidence from European EHIS data suggest that use of any colonoscopy within 10 years may have further increased,^20^ which may explain the even lower incidences for 2016.

### Validity of the Simulated Effects of Colonoscopy Screening

As well, the model was able to approximately reproduce the effect of the successive uptake of colonoscopy utilization in terms of relative changes in incidence patterns over time. The drop in 15-year cumulative incidences with varying levels of annual colonoscopy (0.2-0.6 percent units in men, 0.1-0.5 percent units in women) as predicted by the model was slightly more pronounced, but overall comparable to the change over time calculated for registry-derived estimates before and after the introduction of colonoscopy screening in 2002 (0.1-0.4 percent units in both men and women). This suggests that the modelled relative effect of colonoscopy, though inarguably dependent on appropriately specified input parameters, adequately reproduces effects seen in the real-world.

Additional evidence supporting the validity of the modelled effect of screening colonoscopy comes from the simulation of a hypothetical colonoscopy screening RCT, which yielded outcome patterns comparable to those reported for the sigmoidoscopy screening RCTs. We chose the sigmoidoscopy-RCTs as basis for comparison since it appears reasonable to assume that long-term results from randomized studies comparing colonoscopy versus no screening, which are not to be expected before the late 2020s,^21^ will yield similar patterns. The seen initial increase in cumulative incidence in the intervention group may be expected to be even more pronounced, given that sigmoidoscopy is limited to the rectum and distal part (where approximately 2/3 of CRCs are found), while colonoscopy examines the entire colon.

Accordingly, the potential for relative reduction of the CRC incidence and mortality likely is even higher for colonoscopy as compared to sigmoidoscopy. A 2014 meta-analysis of observational studies on the effect of colonoscopy found an incidence reduction of 69% and a mortality reduction of 68% (**Supplementary Table 7**).^4^ More recent studies found comparable estimates.^8,9^ In the per-protocol analyses of our hypothetical RCT, we found an incidence reduction by screening colonoscopy of approximately 60-65%, with an associated mortality reduction of 75-80%. Other simulation studies found similar-to-larger effects, reaching up to 88% CRC incidence reduction and 90% CRC mortality reduction.^22^ Taken together, our model compares well to the results of the 2014 meta-analysis but may be more conservative than other simulation models, which seems appropriate given the large confidence intervals for the screening colonoscopy effect found in the 2014 meta-analysis.

Finally, our model predicted similar effects of screening colonoscopy in men and women. This seems reasonable as currently, to our knowledge, there is no evidence from colonoscopy studies suggesting otherwise. The evidence from sigmoidoscopy RCTs is ambiguous, indicating a possibly greater (SCORE), smaller (PLCO, UKFSS) or even no (NORCCAP) effect of screening in women. A pooled analysis suggested stronger incidence reduction in younger women as compared to men but no screening effect in older women.^23^ Reasons why sigmoidoscopy screening should have limited or no effect in women are unclear with no plausible rationale at hand,^6^ and chance findings due to underpowered studies by limited sample size for sex-stratified analyses cannot be ruled out.

### Strengths and Limitations

Literature on previous modelling studies on the effect of CRC screening^11,24^ suggests that simulation models are frequently complicated by the need to balance consistent, high-quality and reliable data sources against desired functionality on the one hand, and transparency and reproducibility against conciseness and focus on the other. With this in mind, our motivation was to develop a model explicitly for the screening eligible population, derived from consistent, comprehensible and high-quality data sources strongly linked to the German CRC screening landscape, and reproducible with reasonable efforts.

Our model’s strong reliance on parameters derived from exceptionally large, partly unique real-world data sources (i.e., the German national colonoscopy registry) therefore represents a key conceptual strength. However, the inherent downside of this reliance is that it complicates assessing the model’s external validity, which evidently depends on the existence of data the model ought to reproduce. For instance, a microsimulation model calibrated to the Norwegian population was validated by replicating and predicting outcomes from the (Norwegian) NORCCAP RCT.^25^ A major limitation of this study was that we could not provide a similar validation, as no comparably suitable large-scale trial at the highest level of evidence has been conducted in the German population. Based on similar grounds, we could not assess the model’s ability to reproduce effects of fecal testing in a German population, even though the model can principally be used to simulate stool-test based screening at varying intervals. ^26^

As alternative, we chose the herein presented three-fold approach, which uses the currently best available evidence base to validate a simulation model developed from epidemiological data in Germany. This approach enabled us to scrutinize the model’s natural history component (part 1 and 2) as well as the modelled effect of screening colonoscopy (part 2 and 3) at the same time. Colonoscopy (but not sigmoidoscopy) has been offered as primary screening modality in Germany since 2002, and inarguably constitutes the final pathway of any two-tier screening modality (including the since 1975 offered fecal stool tests) for the foreseeable future.^11^

In part 1 of our validation approach, a limitation was that the outcome data of screening colonoscopy participants collected in the KolosSal study are partly also documented in the National Screening Colonoscopy Registry, which was used to derive the transition rates between model states. However, the subsample of KolosSal participants from 2005-2012 (N = 11,912) was only a minute fraction (approximately 0.3%) of the Colonoscopy Registry data (N = 3.6 to 4.3 million), and excluding KolosSal participants recruited in 2005-2012 from the Colonoscopy Registry data which would have insured complete independence would not have changed the model parameters to any relevant extent.

Furthermore, part 2 of the validation study is limited to comparisons of annual colonoscopy use of 2% and 6%, assumed to be representative for the years 2001 and 2011, respectively. We refrained from simulating a scenario for 2016 due to the lack of reliable input parameters. Apart from the annual colonoscopy use, this also includes potential previous screening. As screening colonoscopy was introduced in 2002 and is typically recommended at 10-yearly intervals, considerable – but unknown – proportion of subjects will likely have undergone a repeated screening colonoscopy from 2011 onwards.

Finally, no distinction according to cancer subsite can be made by the model, which would have allowed to separately scrutinize the effect of endoscopic screening on distal cancers in part 3 of the validation. While the German national screening colonoscopy registry was a particularly well-suited data source to derive key model parameters, the registry did not include sufficiently detailed data to calculate specific transition rates for proximal and distal neoplasms.

## Conclusions

Despite these limitations, this study suggests that the model may adequately predict colorectal neoplasm prevalences and incidences in a German population for up to 25 years, with estimated patterns of the effect of screening colonoscopy resembling those seen in registry data and real-world studies. The model may therefore be valid to assess the comparative effectiveness of strategies for colorectal cancer screening, and thus represent powerful and transparent tool to inform medical decision making.

## Declarations

### Ethics approval

The KoloSsal study was approved by ethics committees of the University of Heidelberg (057/2005) and of the Medical Association of Saarland (54/05).

### Consent to participate and for publication

Informed consent was obtained from each participant.

### Data availability statement

All analyses relevant to the study are included in the article or uploaded as supplementary information. The model source code developed in the statistical software R is uploaded as supplementary information. All data requests regarding the KolosSal study should be submitted to the corresponding author for consideration. Access to anonymised data may be granted following review.

## Data Availability

All analyses relevant to the study are included in the article or uploaded as supplementary information. The R code defining the core model is uploaded as supplementary information. All data requests regarding the KolosSal study should be submitted to the corresponding author for consideration. Access to anonymised data may be granted following review.

## Authors’ contributions

HB and TH designed this study and developed the methodology. HB designed, initiated and led the KolosSal study. MH and HB contributed to the acquisition of data. TH conducted the statistical analyses and drafted the manuscript. All authors critically reviewed the manuscript, contributed to its revision, and approved the final version submitted. The researchers are independent from funders. All authors had full access to all of the data (including statistical reports and tables) used for the study and can take responsibility for the integrity of the data and the accuracy of the data analysis.

## Acknowledgements

The authors thank all individuals who participated in the KolosSal study, all participating gastroenterologists for the recruitment of study participants, and all staff involved in study coordination, documentation and data preparation

## Preprint

doi.org/10.1101/2020.04.17.20069484

## Abbreviations

ADN: any advanced neoplasm
ANN: any neoplasm
CI: confidence interval
CRC: colorectal cancer
ITT: intention to treat
KOLOSSAL: Effektivität der Früherkennungs-Koloskopie: eine Saarland-weite Studie
NORCCAP: Norwegian Colorectal Cancer Prevention trial
PLCO: Prostate, Lung, Colorectal and Ovarian trial PP per protocol
PRR: prevalence ratio
RCT: randomized controlled trial
SCORE: Screening for Colon Rectum trial
TOST: two one-sided t-tests
UKFSS: United Kingdom Flexible Sigmoidoscopy Screening

