## Supplementary Appendix 1c for "Effects of Screening for Colorectal Cancer: Development, Documentation and Validation of a Multistate Markov Model"

Modeling Colorectal Cancer Screening

Thomas Heisser, Michael Hoffmeister, Hermann Brenner

**Supplementary Appendix 1a: Model Documentation.**

**Supplementary Appendix 1b: R Code.**

**Supplementary Appendix 1c: Data Sources and Statistical Analysis.**

**Supplementary Appendix 2: Supplementary Tables and Figures.**

### **Supplementary Appendix 1c. Data Sources and Statistical Analysis**

### **Data Sources**

#### KolosSal Study

Details on the KolosSal study have been described previously.^1–5^ Briefly, the KolosSal study recruited patients undergoing screening colonoscopy with the objective to monitor long-term CRC incidence and mortality. Residents of the federal state of Saarland aged 55 or older undergoing screening colonoscopy in a participating practice were eligible for enrolment. The study was approved by ethics committees of the University of Heidelberg and of the State Medical Association of Saarland. Informed consent was obtained from each patient.

This analysis is based on data of participants recruited between 2005 and 2013 in 33 gastroenterological practices in Saarland certified to perform screening colonoscopy. Patients were classified according to the most advanced of the following findings: CRC, advanced adenoma – defined as presence of at least 1 adenoma with at least 1 of the following features: >1cm in size, tubulovillous or villous components, high-grade dysplasia – other adenoma, hyperplastic or unspecified polyp, none of these.^6,7^

#### German Registry Data

Age-group- and sex-specific incidence rates for the German general population were obtained from the interactive database of the German Centre for Cancer Registry Data,^8^ which provides combined and quality-checked data from epidemiological cancer registries in Germany. We extracted the CRC incidence rates for the years 2001, 2006, 2011, and 2016, i.e. one year before the introduction of the population-based colonoscopy screening offer in Germany in 2002, and then in steps of five years to allow for detecting effects of the successive uptake of the colonoscopy screening offer (**Supplementary Table 4**).^9^

### **Statistical Analysis**

#### Part 1: Equivalence to KolosSal Study Prevalences

Of the KolosSal study population, we selected subjects aged 55-75 who had their first screening colonoscopy. We excluded patients with history of inflammatory bowel disease and patients where no information could be obtained on whether a previous colonoscopy had been conducted. We then assessed the detected prevalences of any neoplasm (ANN, any adenomas and cancers) and of any advanced neoplasm (ADN, advanced adenomas and cancer) at screening colonoscopy according to age at screening in steps of one year from age 55-75, stratified by sex, and calculated the 95% confidence intervals (CI).

We then modelled, for a hypothetical population of previously unscreened men and women, disease progression without screening from the states at ages 50-75. For each age group, the simulated number of subjects was matched to the number of subjects as included from the KolosSal study population. We assessed the prevalences of ANN and ADN which would have been detected if screening colonoscopy had been applied once at each individual age from ages 55-75, and calculated the 95% CIs.

Subsequently, we calculated the prevalence ratios (PRR) by dividing the modelled prevalence estimates by the prevalences observed in KolosSal, both age-specific and as multilevel meta-analyses combining all sex-specific PRRs, along with the respective CIs. To account for potential dependencies in the data, we defined age group as additional level in the meta-analysis. Statistical equivalence between the modelled and observed prevalences was then tested by applying two one-sided t-tests (TOST)^10^ to each of the sex-specific meta-analyses estimates. We chose a 20% margin of equivalence. A maximum deviation of 20% was considered acceptable as the model is not primarily intended to meticulously forecast absolute case numbers with or without screening but rather to assess the comparative performance of CRC screening strategies in terms of their contribution to the relative reduction of the CRC burden.^11^

#### Part 2: Comparison to Registry-derived Cumulative Incidences

For each of the extracted years (2001, 2006, 2011, 2016) from the database of the German Centre for Cancer Registry Data, we estimated the 15-year cumulative CRC incidence for men and women of the German general population aged 50-64, 55-69 and 60-74. As the incidence rates as provided by the registry are computed for age intervals of five years, the 15-year cumulative rate is calculated as sum of five times the age-group-specific interval rate for the respective age-groups, assuming the age-specific rates remain constant over time. The cumulative risk in the absence of competing causes of death can then be approximated by the cumulative rate.^12,13^

For comparison, we derived corresponding estimates for modelled populations of each 100,000 German men and women with levels of 2% and 6% annual colonoscopy use. These utilization levels were chosen as the estimated use of colonoscopy for any reason within 10 years in Germany was 24.3% and 63.9% for the periods 2000-2002 and 2008-2010, respectively, reflecting the increase triggered by the introduction of the colonoscopy screening offer in 2002.^9^

As the model starting prevalences were obtained from a population of first-time screening colonoscopy users,^14^ i.e. a selective population without or only negative prior fecal testing or (diagnostic) colonoscopy, we discounted adenoma and preclinical cancer prevalences at model start by 30%. This was considered reasonable in order to increase comparability to the German general population given that, already in the period 2000-2002, approximately 66% of 55-59 year-olds had either fecal testing within two years or colonoscopy within ten years.^9^

#### Part 3: Comparison to RCT-reported CRC Incidence and Mortality Patterns

Finally, we modelled each 100,000 50- and 60-year old men and women with and without screening colonoscopy at baseline with 16 years of follow-up, i.e. the approximate median follow-up length of by now reported results from screening-sigmoidoscopy RCTs.^15–18^ We assumed 70% adherence to baseline colonoscopy in the screening group and 30% (additional) use of colonoscopy over the course of the follow-up period in both groups to account for colonoscopy utilization rates in the general population regardless of indication.^19^ Between both groups, we compared the cumulative incidence and mortality after the end of follow-up and the annual incidence and mortality rate ratio over time. The thereof resulting patterns of the effect of screening colonoscopy were compared to the patterns reported in screening sigmoidoscopy RCTs.
