## Supplementary Appendix 2 for "Effects of Screening for Colorectal Cancer: Development, Documentation and Validation of a Multistate Markov Model"

Modeling Colorectal Cancer Screening

Thomas Heisser, Michael Hoffmeister, Hermann Brenner

**Supplementary Appendix 1a: Model Documentation.**

**Supplementary Appendix 1b: R Code.**

**Supplementary Appendix 1c: Data Sources and Statistical Analysis.**

**Supplementary Appendix 2: Supplementary Tables and Figures.**

### **Supplementary Appendix 2.**

**Overview**

**Supplementary Table 4**. Sex- and age-specific incidence rate as extracted from the database of the German Centre for Cancer Registry Data

**Supplementary Table 5**. Characteristics of the KolosSal study population included in the analyses

**Supplementary Table 6.** Sensitivity Analysis (95% prediction bands). Registry- vs. model-derived cumulative incidence after 15 years for ages 50-64, 55-69, and 60-74.

**Supplementary Table 7**. Comparison of effects of screening colonoscopy seen in the model to those reported in a meta-analysis of observational studies

**Supplementary Figure 1**. Testing for equivalence between model and real study outcomes by applying two one-sided t-tests (TOST).

**Supplementary Figure 2**. Numbers needed to screen to prevent one event (diagnosis of colorectal cancer or colorectal cancer death) after 15 simulated years

**Supplementary Figure 3.** Cumulative Incidence and Mortality Patterns in SCORE, NORRCAP, PLCO and UKFSS

**Supplementary Figure 4**. Annual incidence and mortality rate ratios (screening /control).

**Supplementary Figure 5.** Cumulative incidence at different starting ages of the simulation, stratified by sex.

**Supplementary Figure 6.** Cumulative mortality at different starting ages of the simulation, stratified by sex.

**Supplementary Figure 7.** Annual incidence rate ratios (screening /control), stratified by sex.

**Supplementary Figure 8**. Annual mortality rate ratios (screening /control), stratified by sex.

#### **Supplementary Table 4.** Sex- and age-specific incidence rate as extracted from the database of the German Centre for Cancer Registry Data^1^

|  | **Crude incidence rate per 100,000 inhabitants** | | | | | | | | |
| --- | --- | --- | --- | --- | --- | --- | --- | --- | --- |
| **Age Group** | **Men** | | | |  | **Women** | | | |
|  | **2001** | **2006** | **2011** | **2016** |  | **2001** | **2006** | **2011** | **2016** |
| 50 - 54 | 62.6 | 52.0 | 53.3 | 47.7 |  | 43.4 | 42.8 | 38.9 | 34.5 |
| 55 - 59 | 106.9 | 115.1 | 100.6 | 87.9 |  | 71.2 | 70.0 | 62.9 | 55.7 |
| 60 - 64 | 180.2 | 182.6 | 166.6 | 141.4 |  | 100.4 | 111.1 | 95.3 | 75.4 |
| 65 - 69 | 254.2 | 252.2 | 235.6 | 207.0 |  | 146.8 | 142.4 | 127.3 | 116.0 |
| 70 - 74 | 368.4 | 356.7 | 309.3 | 272.7 |  | 223.3 | 210.3 | 176.5 | 151.7 |
| 75 - 79 | 444.2 | 460.7 | 401.6 | 331.0 |  | 300.5 | 276.8 | 235.1 | 204.8 |
| 80 - 84 | 538.3 | 527.6 | 478.5 | 400.5 |  | 402.0 | 368.5 | 320.6 | 264.0 |

^1^Extracted from reference ^1^

#### **Supplementary Table 5.** Characteristics of the KolosSal study population included in the analyses

|  | Men | |  | | Women | | |
| --- | --- | --- | --- | --- | --- | --- | --- |
| Characteristic | n | % |  | | n | | % |
| Total | 5916 | - | |  | | 5996 | - |
| Age (*y*) |  |  | |  | |  |  |
| 55 | 715 | 12.1% | |  | | 803 | 13.4% |
| 56 | 573 | 9.7% | |  | | 650 | 10.8% |
| 57 | 442 | 7.5% | |  | | 478 | 8.0% |
| 58 | 370 | 6.3% | |  | | 418 | 7.0% |
| 59 | 332 | 5.6% | |  | | 346 | 5.8% |
| 60 | 308 | 5.2% | |  | | 308 | 5.1% |
| 61 | 290 | 4.9% | |  | | 285 | 4.8% |
| 62 | 250 | 4.2% | |  | | 252 | 4.2% |
| 63 | 260 | 4.4% | |  | | 232 | 3.9% |
| 64 | 240 | 4.1% | |  | | 239 | 4.0% |
| 65 | 252 | 4.3% | |  | | 232 | 3.9% |
| 66 | 251 | 4.2% | |  | | 264 | 4.4% |
| 67 | 252 | 4.3% | |  | | 240 | 4.0% |
| 68 | 253 | 4.3% | |  | | 220 | 3.7% |
| 69 | 229 | 3.9% | |  | | 214 | 3.6% |
| 70 | 202 | 3.4% | |  | | 180 | 3.0% |
| 71 | 190 | 3.2% | |  | | 179 | 3.0% |
| 72 | 159 | 2.7% | |  | | 155 | 2.6% |
| 73 | 137 | 2.3% | |  | | 126 | 2.1% |
| 74 | 112 | 1.9% | |  | | 95 | 1.6% |
| 75 | 99 | 1.7% | |  | | 80 | 1.3% |
| School education (*y*) |  |  | |  | |  |  |
| <10 | 3577 | 61.4% | |  | | 3975 | 67.0% |
| 10+ | 2253 | 38.6% | |  | | 1957 | 33.0% |
| Family history of   colorectal cancer |  |  | |  | |  |  |
| No | 5257 | 88.9% | |  | | 5284 | 88.1% |
| Yes | 659 | 11.1% | |  | | 712 | 11.9% |
| Smoking |  |  | |  | |  |  |
| Never | 2159 | 36.9% | |  | | 3555 | 59.7% |
| Current | 705 | 12.0% | |  | | 734 | 12.3% |
| Former | 2988 | 51.1% | |  | | 1664 | 28.0% |
| Body mass index (kg/m^2^) |  |  | |  | |  |  |
| <25 | 1519 | 25.7% | |  | | 2373 | 39.6% |
| 25 – 29.9 | 3018 | 51.0% | |  | | 2280 | 38.0% |
| ≥ 30 | 1379 | 23.3% | |  | | 1343 | 22.4% |

*^a^* Numbers do not add up to the total numbers because of missing values for some variables (in parentheses, men/women): school education (86/64), smoking (64/43)

#### **Supplementary Table 6.** Sensitivity Analysis (95% prediction intervals derived by probabilistic sensitivity analysis in 1000 runs). Registry*- vs. model**-derived cumulative incidence of colorectal cancer after 15 years from baseline for ages 50-64, 55-69, and 60-74

*2.5% Quantile (Lower Limit)*

|  | **Men** | | | | | | |  | **Women** | | | | | | |
| --- | --- | --- | --- | --- | --- | --- | --- | --- | --- | --- | --- | --- | --- | --- | --- |
|  | **CRC Risk (%)** | | | | | | |  | **CRC Risk (%)** | | | | | | |
|  | **German Registry** | | | |  | **Model** | |  | **German Registry** | | | |  | **Model** | |
|  | **year** | | | |  | **annual colonoscopy use** | |  | **year** | | | |  | **annual colonoscopy use** | |
| **Age at baseline** | **2001** | **2006** | **2011** | **2016** |  | **2%** | **6%** |  | **2001** | **2006** | **2011** | **2016** |  | **2%** | **6%** |
| 50 | 1.7 | 1.7 | 1.6 | 1.4 |  | 1.4 | 1.2 |  | 1.1 | 1.1 | 1.0 | 0.8 |  | 0.8 | 0.7 |
| 55 | 2.7 | 2.7 | 2.5 | 2.2 |  | 2.4 | 2.1 |  | 1.6 | 1.6 | 1.4 | 1.2 |  | 1.5 | 1.2 |
| 60 | 4.0 | 4.0 | 3.6 | 3.1 |  | 3.9 | 3.3 |  | 2.4 | 2.3 | 2.0 | 1.7 |  | 2.5 | 2.1 |

*97.5% Quantile (Upper Limit)*

|  | **Men** | | | | | | |  | **Women** | | | | | | |
| --- | --- | --- | --- | --- | --- | --- | --- | --- | --- | --- | --- | --- | --- | --- | --- |
|  | **CRC Risk (%)** | | | | | | |  | **CRC Risk (%)** | | | | | | |
|  | **German Registry** | | | |  | **Model** | |  | **German Registry** | | | |  | **Model** | |
|  | **year** | | | |  | **annual colonoscopy use** | |  | **year** | | | |  | **annual colonoscopy use** | |
| **Age at baseline** | **2001** | **2006** | **2011** | **2016** |  | **2%** | **6%** |  | **2001** | **2006** | **2011** | **2016** |  | **2%** | **6%** |
| 50 | 1.7 | 1.7 | 1.6 | 1.4 |  | 2.0 | 1.7 |  | 1.1 | 1.1 | 1.0 | 0.8 |  | 1.2 | 1.0 |
| 55 | 2.7 | 2.7 | 2.5 | 2.2 |  | 3.2 | 2.7 |  | 1.6 | 1.6 | 1.4 | 1.2 |  | 1.9 | 1.6 |
| 60 | 4.0 | 4.0 | 3.6 | 3.1 |  | 4.7 | 4.1 |  | 2.4 | 2.3 | 2.0 | 1.7 |  | 3.0 | 2.5 |

**Registry-derived cumulative incidence was estimated based on age-group and sex-specific incidence rates extracted one year before the introduction of the screening colonoscopy offer in Germany in 2002 and then in steps of 5-years.*

*** Model-derived cumulative incidence assumes approximate levels of annual colonoscopy use before (2%) and after (6%) the introduction of the colonoscopy screening offer.*

*CRC, colorectal cancer.*

#### **Supplementary Table 7.** Comparison of effects of screening colonoscopy seen in the model to those reported in a meta-analysis of observational studies

|  | **Effect of screening colonoscopy at baseline**  **(per protocol)** | | **Meta-Analysis Brenner 2014**^2^ | |
| --- | --- | --- | --- | --- |
| **Age** | **Incidence Reduction (%)** | **Mortality Reduction (%)** | **Incidence Reduction (%, 95% CI)** | **Mortality Reduction (%, 95% CI)** |
| 50 | 61 | 75 | 69 (23 - 88) | 68 (57 - 77) |
| 60 | 64 | 79 |  |  |

*CI, confidence interval.*

#### **Supplementary Figure 1.** Testing for equivalence between model and real study outcomes by applying two one-sided t-tests (TOST). Shown are the meta-analyses estimates and 90% confidence intervals* for the summarized prevalence ratios at ages 55-75 for any neoplasm and any advanced neoplasm. Only the higher of the two p-values for the TOST are shown, values <0.05 suggest statistical equivalence at a 20% margin.

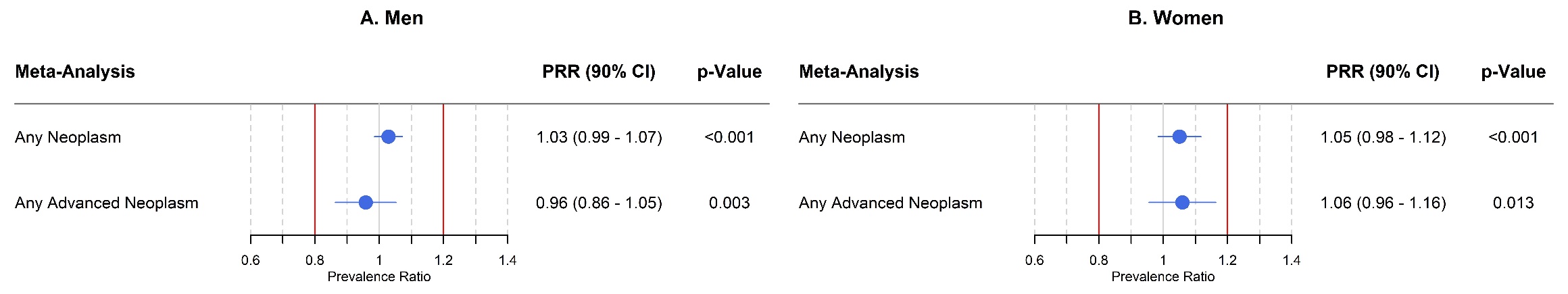

**Using a 90% confidence interval yields a 0*.*05 significance level for testing equivalence (as two one-sided tests without adjustment for multiple testing are performed))*

*PRR, Prevalence Ratio*

#### **Supplementary Figure 2.** Numbers needed to screen* to prevent one event (diagnosis of colorectal cancer or colorectal cancer death) after 15 simulated years, stratified by age at start of the simulation and by sex.

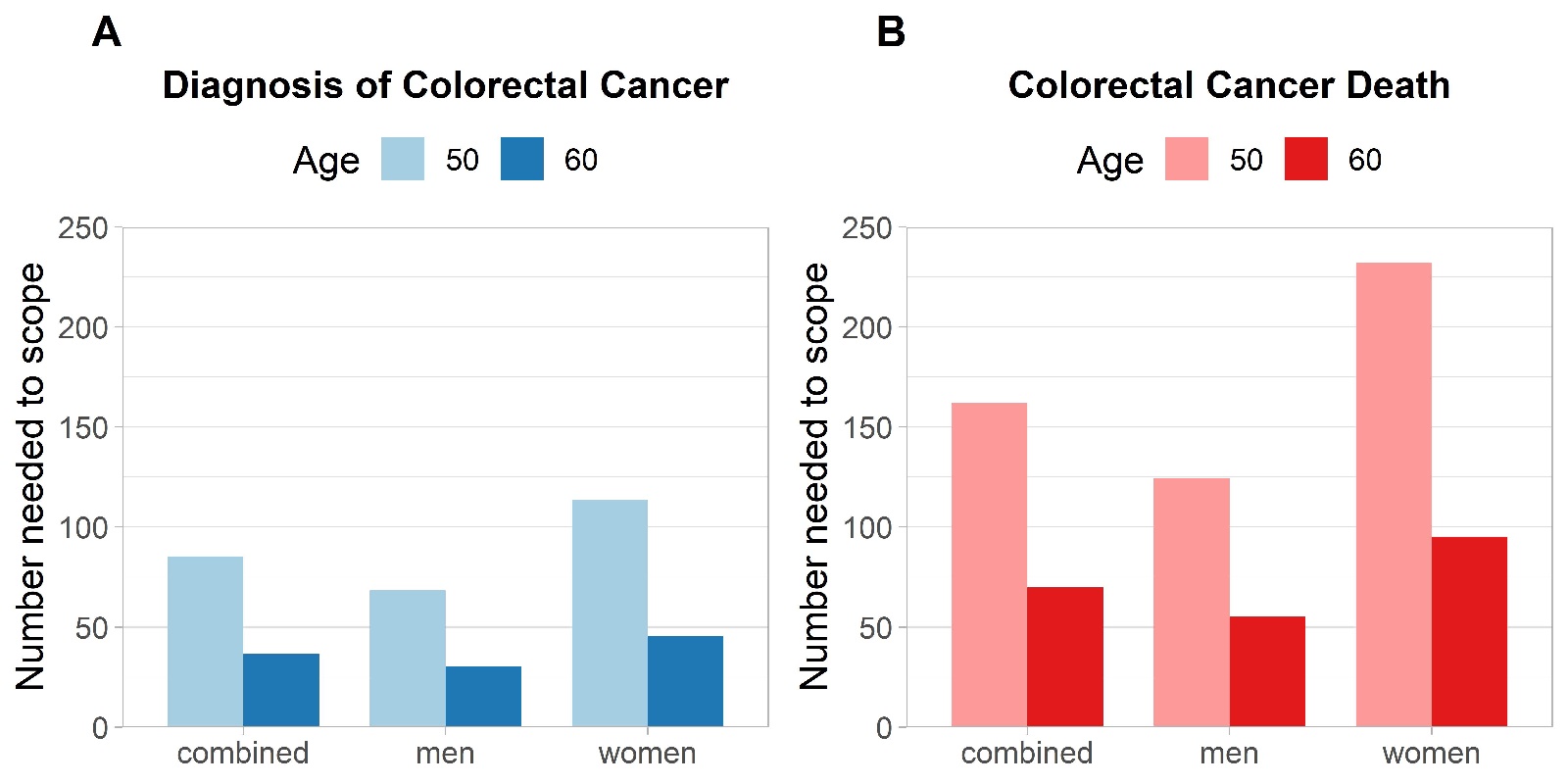

******* *Calculated by dividing the number screened by the number of events prevented in the intervention group.*

#### **Supplementary Figure 3.** Cumulative Incidence and Mortality Patterns in SCORE, NORRCAP, PLCO and UKFSS

**Per Protocol**

| **Cumulative Incidence** | **Cumulative Mortality** |
| --- | --- |
| **SCORE^1^** | |
| 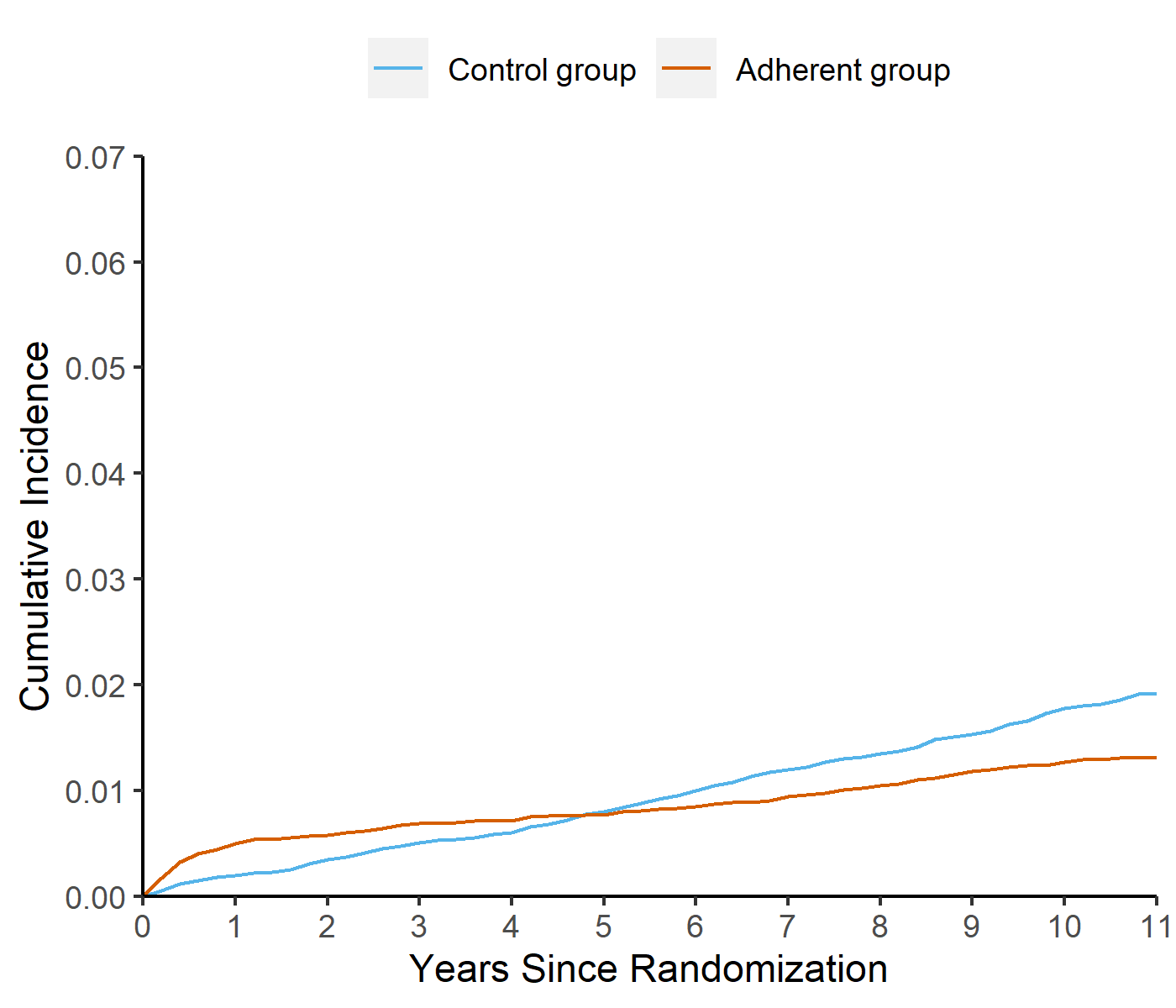 | 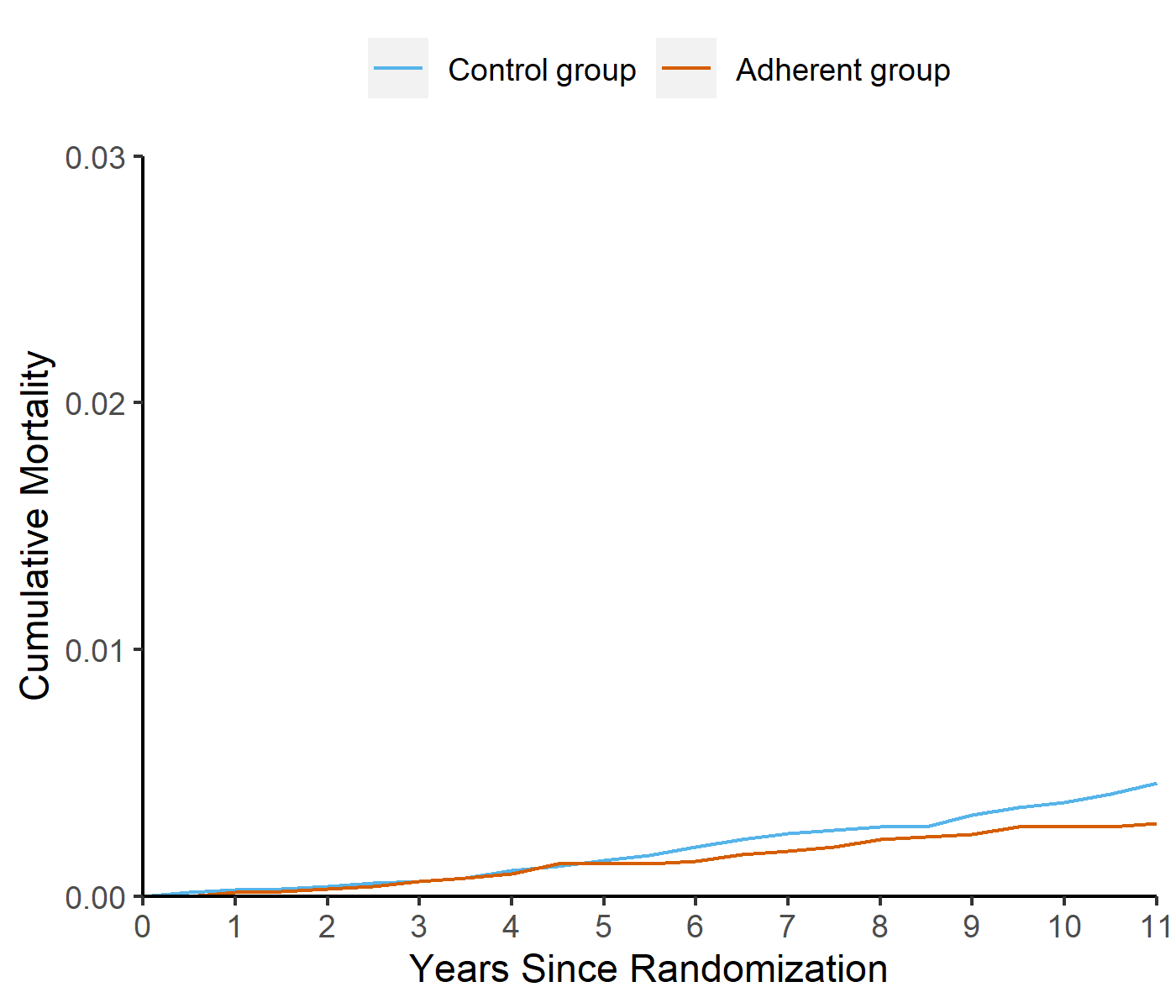 |
| **NORCCAP^2^** | |
| 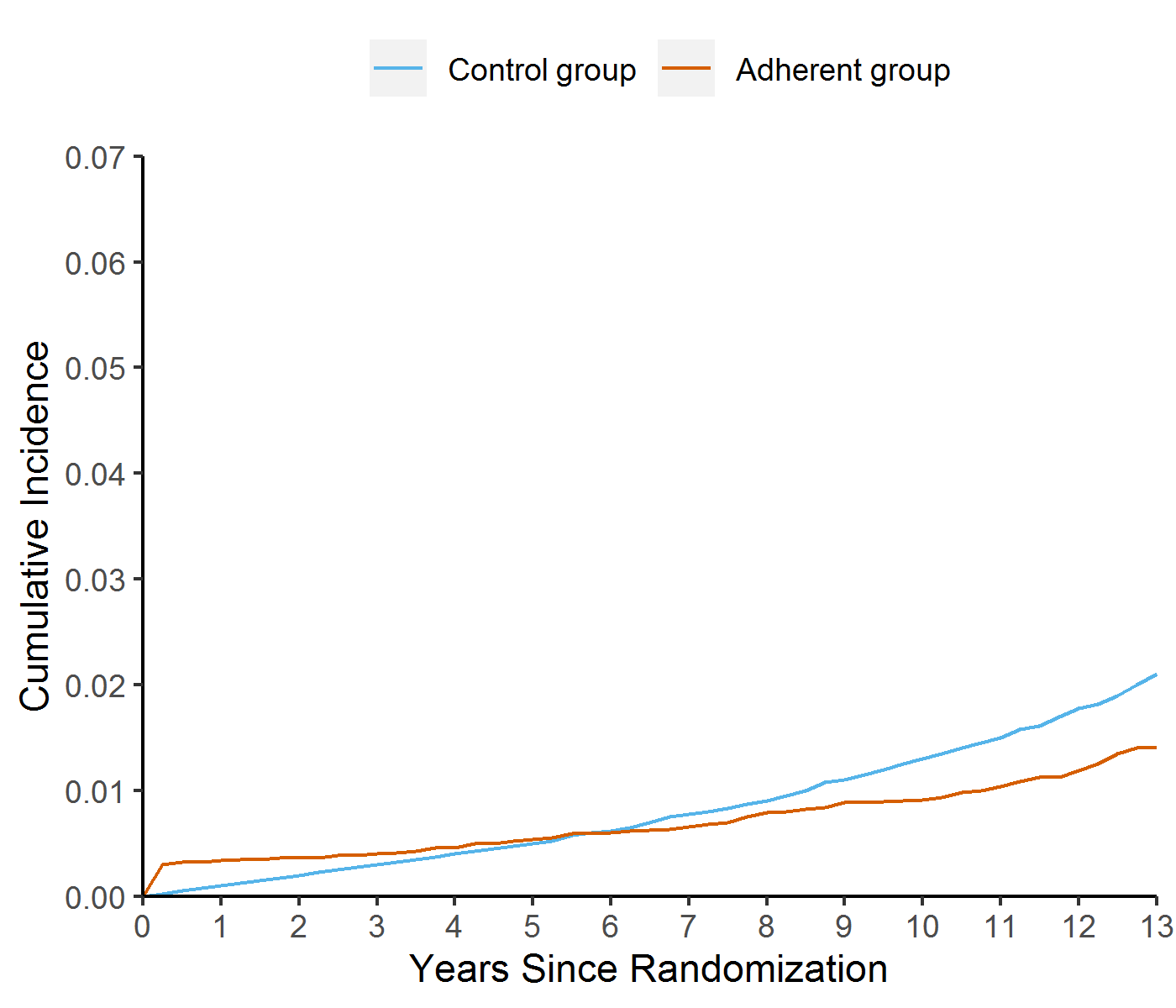 | 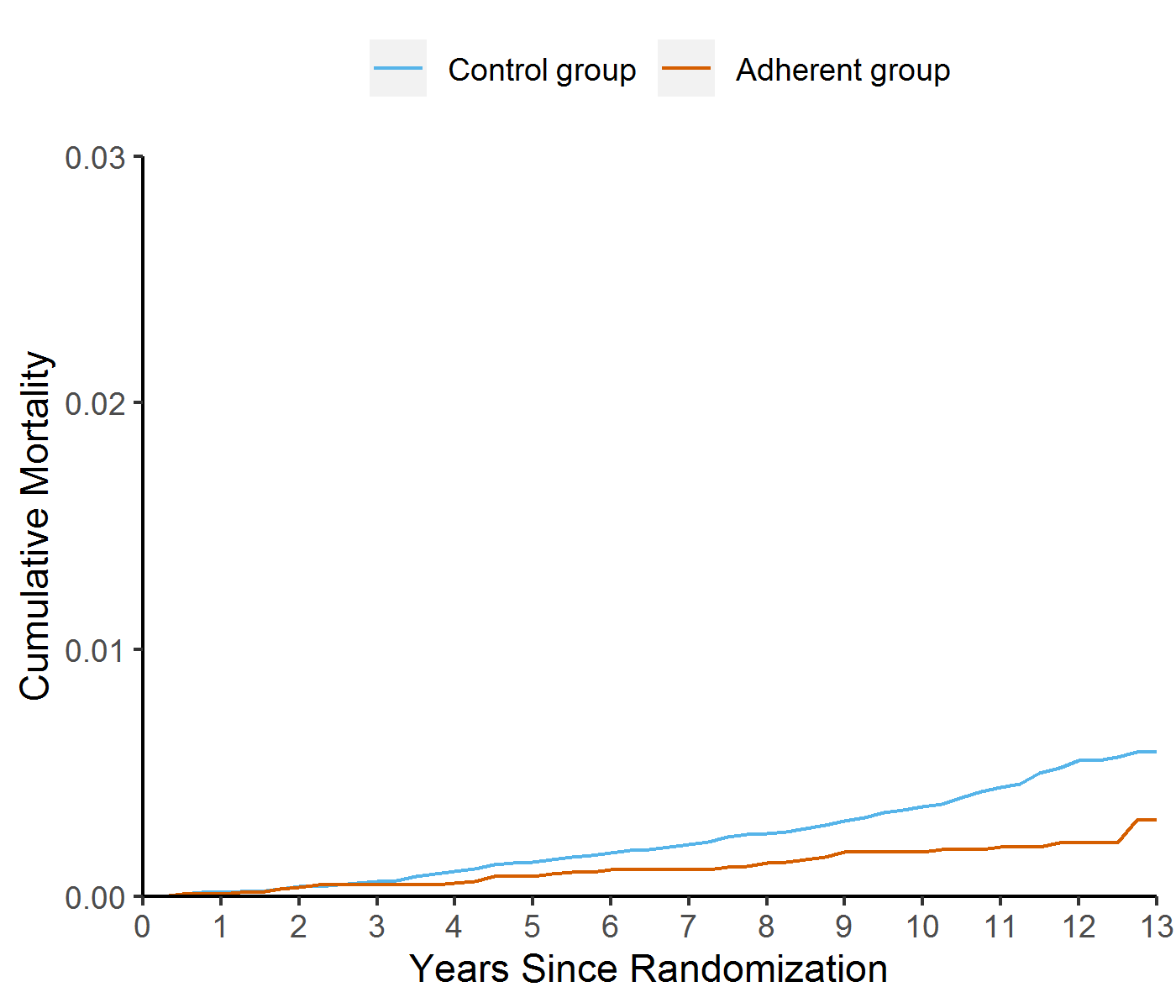 |
| **UKFSS^3^** | |
| 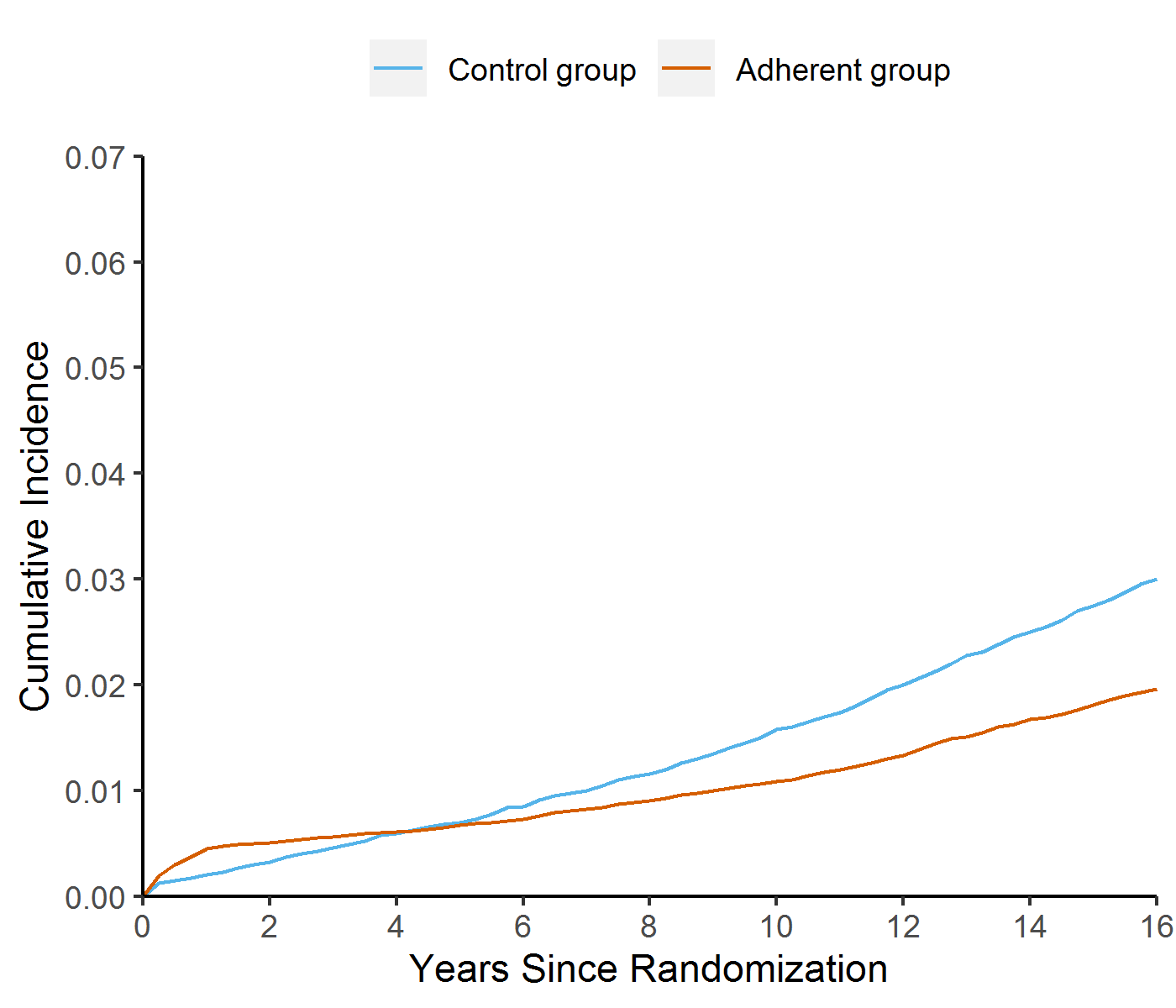 | 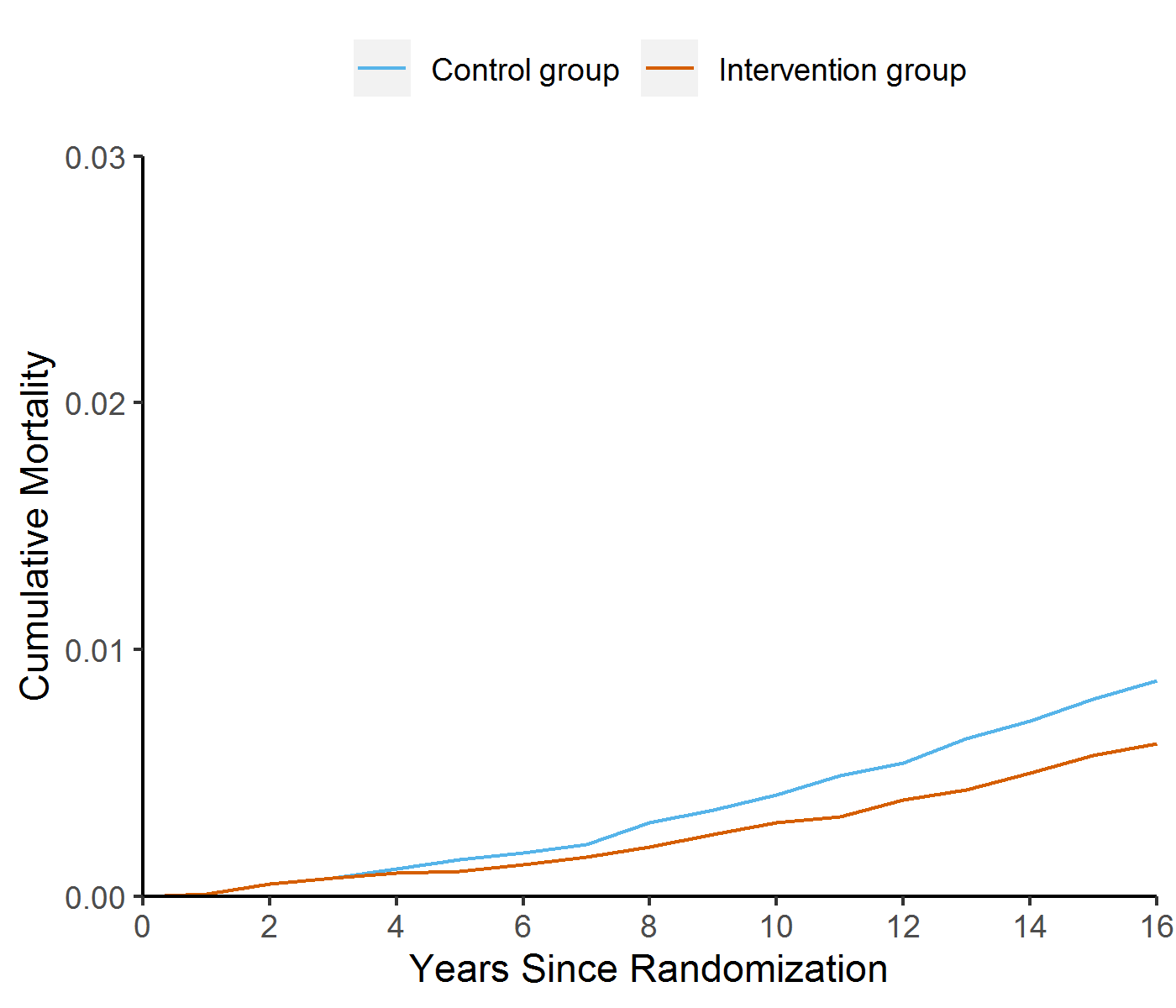 |
| **PLCO** | |
| *Not available* | *Not available* |

**Intention to Screen**

| **Cumulative Incidence** | **Cumulative Mortality** |
| --- | --- |
| **SCORE^1^** | |
| 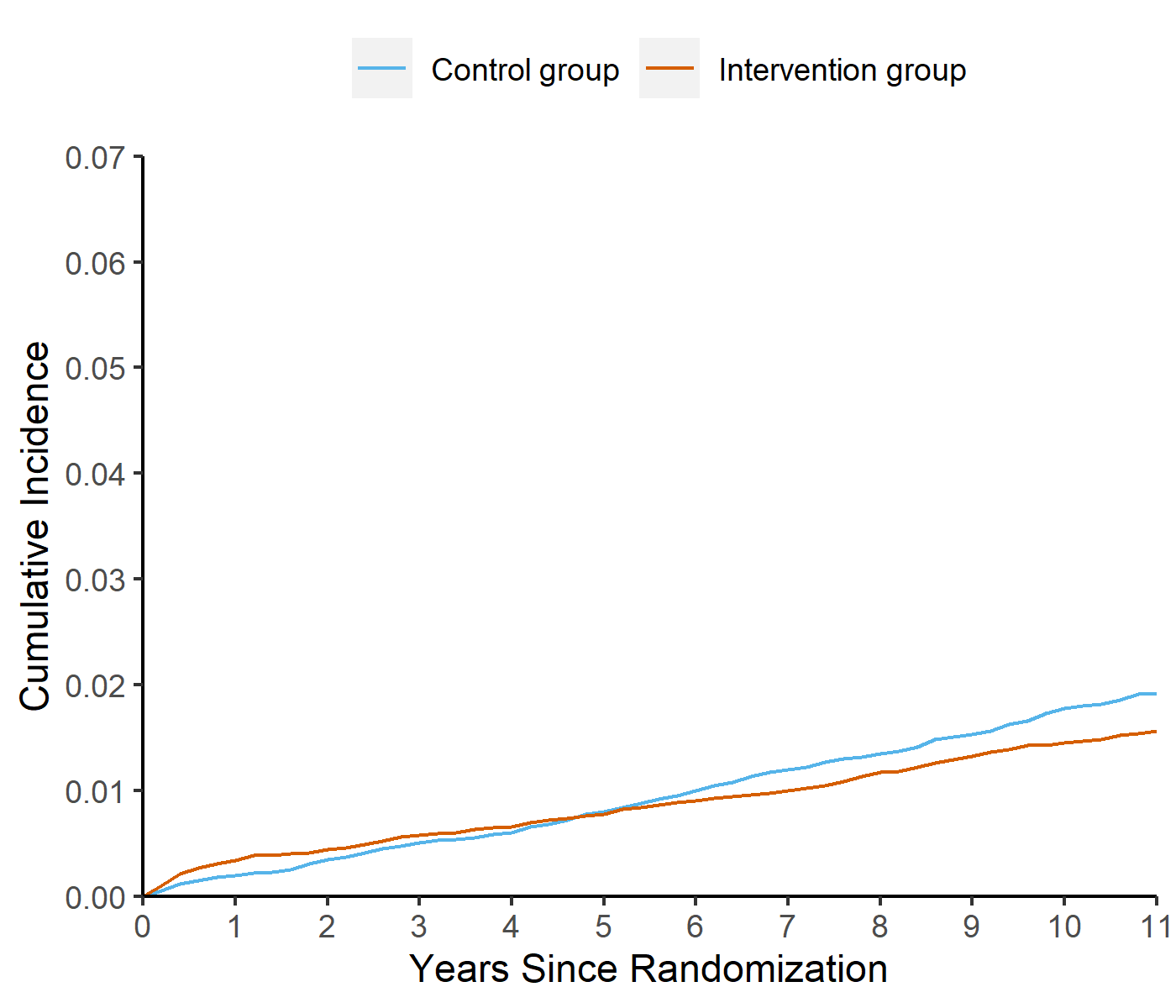 | 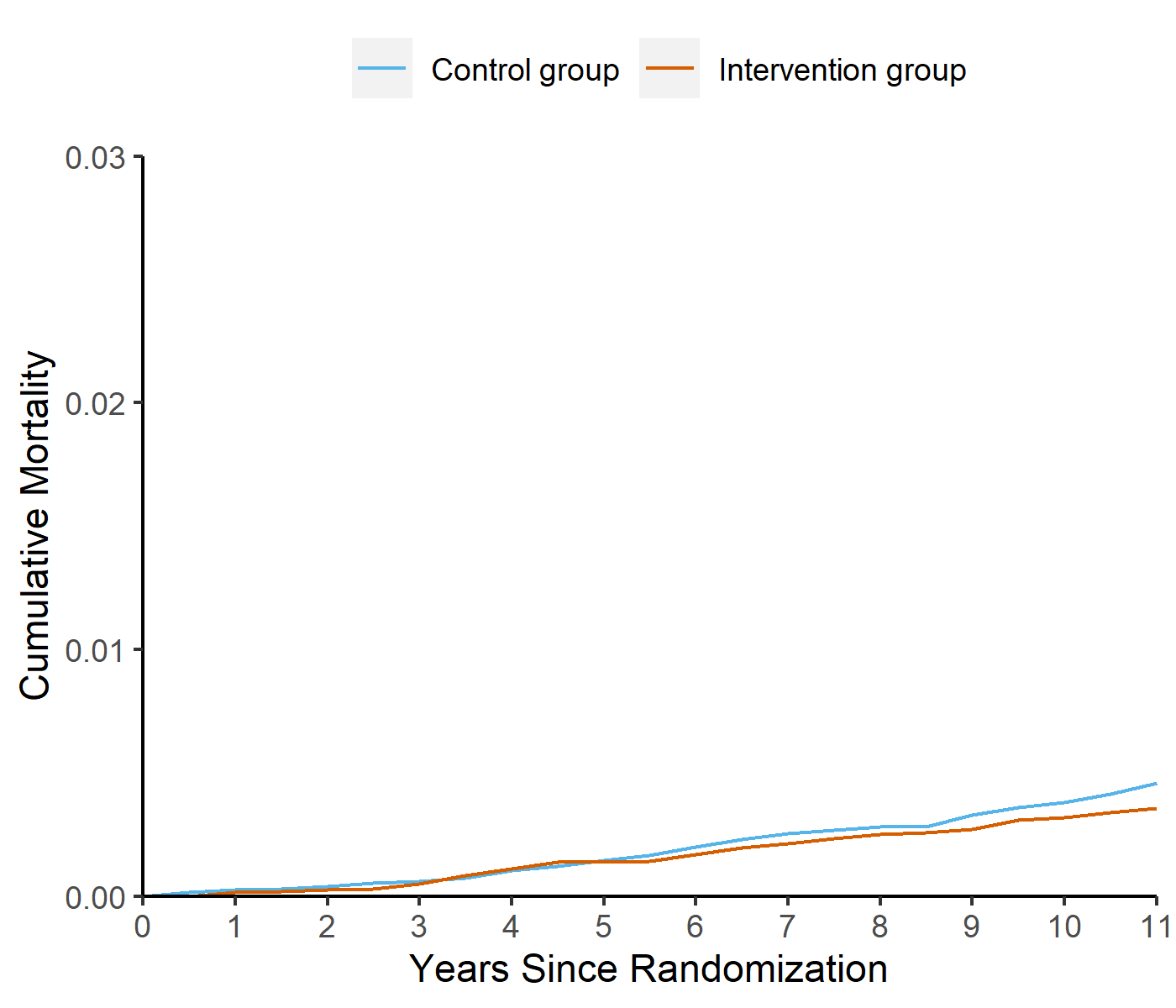 |
| **NORCCAP^2^** | |
| 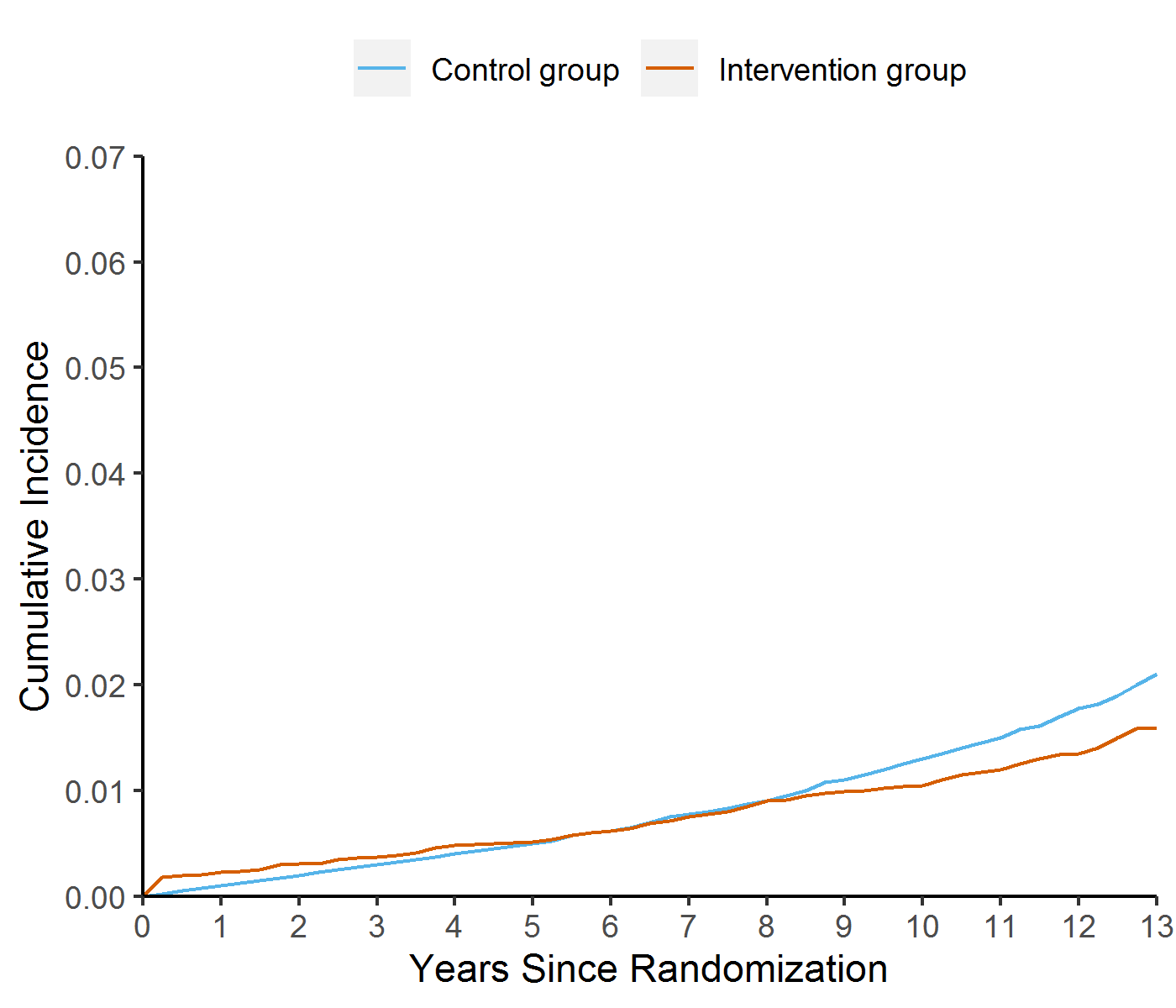 | 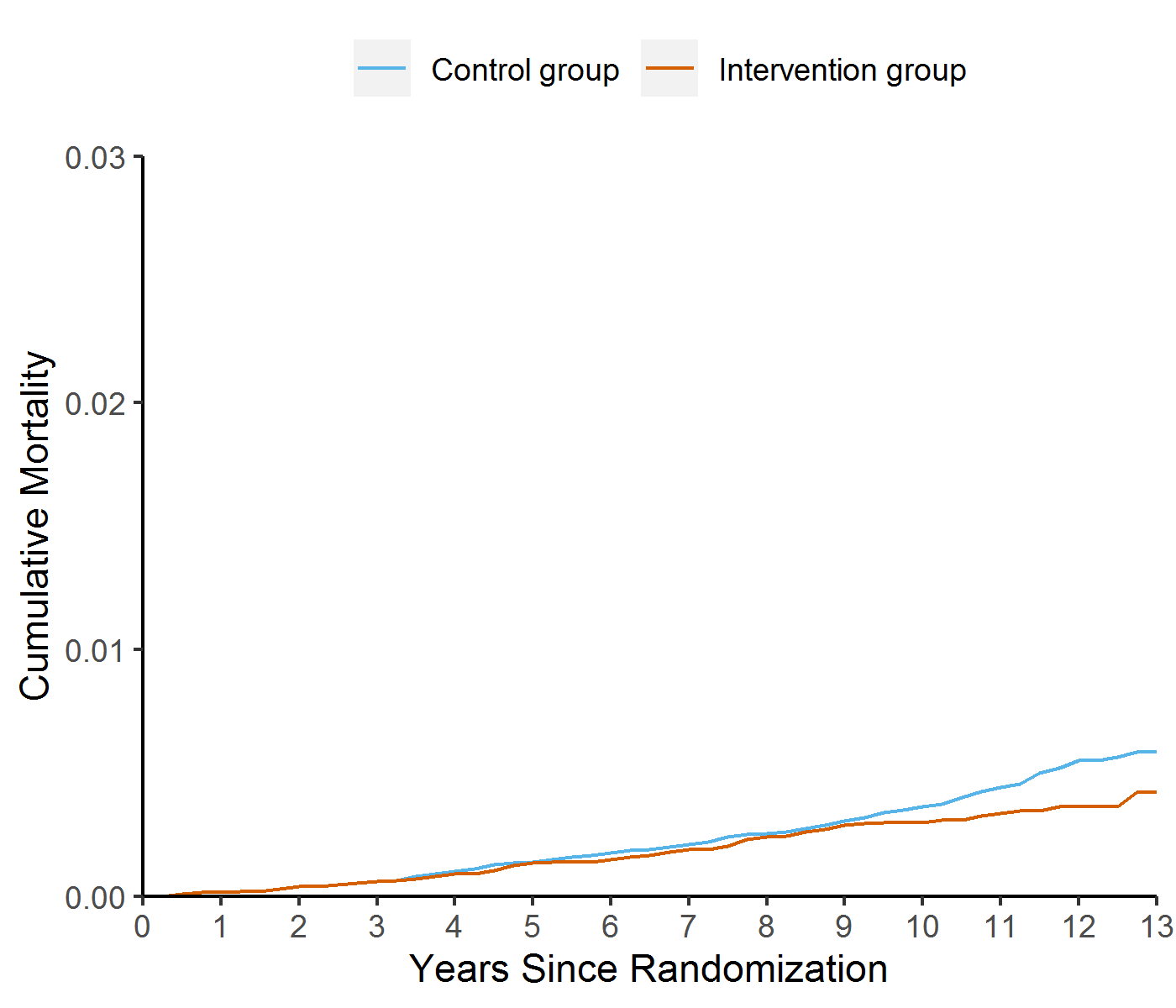 |
| **UKFSS^3^** | |
| 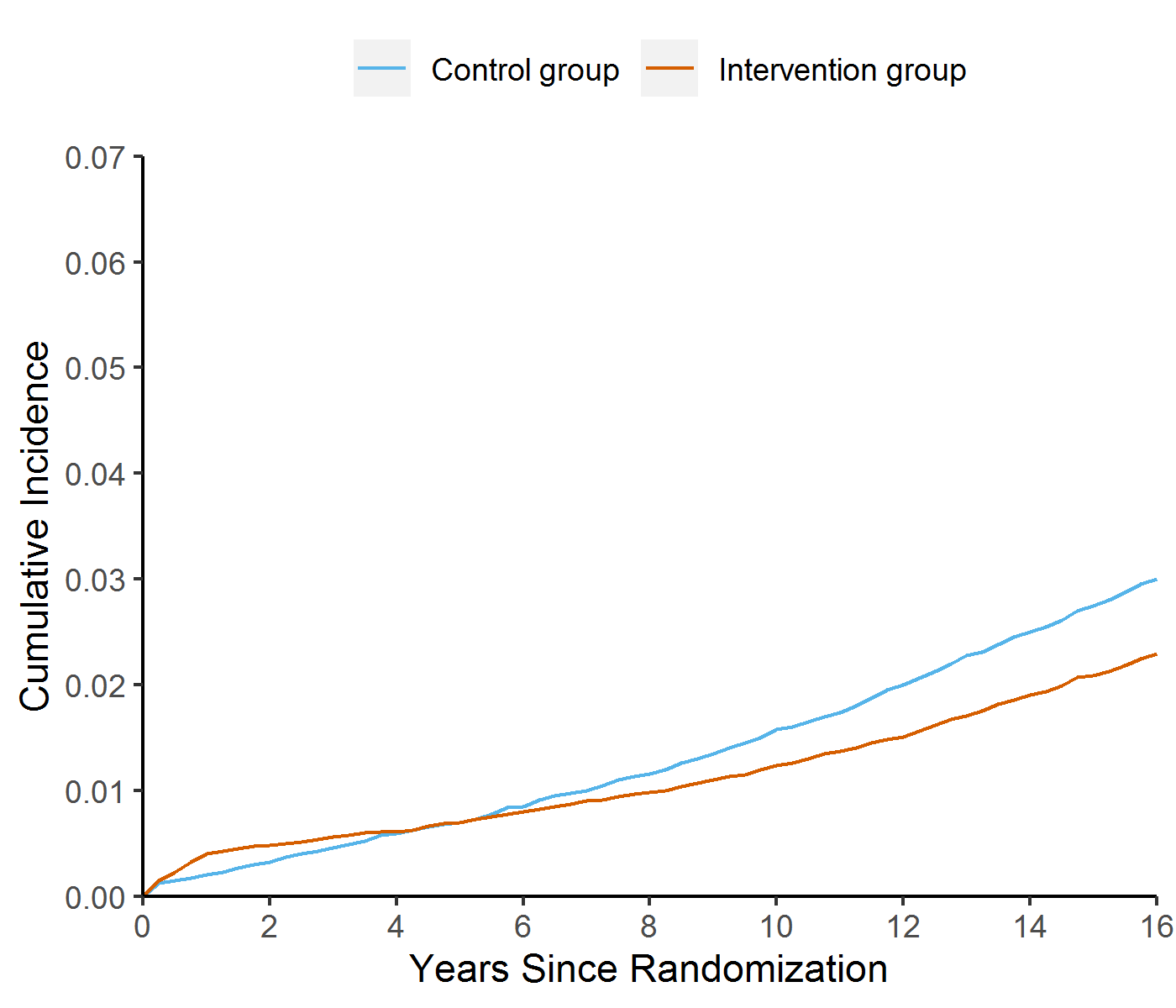 | 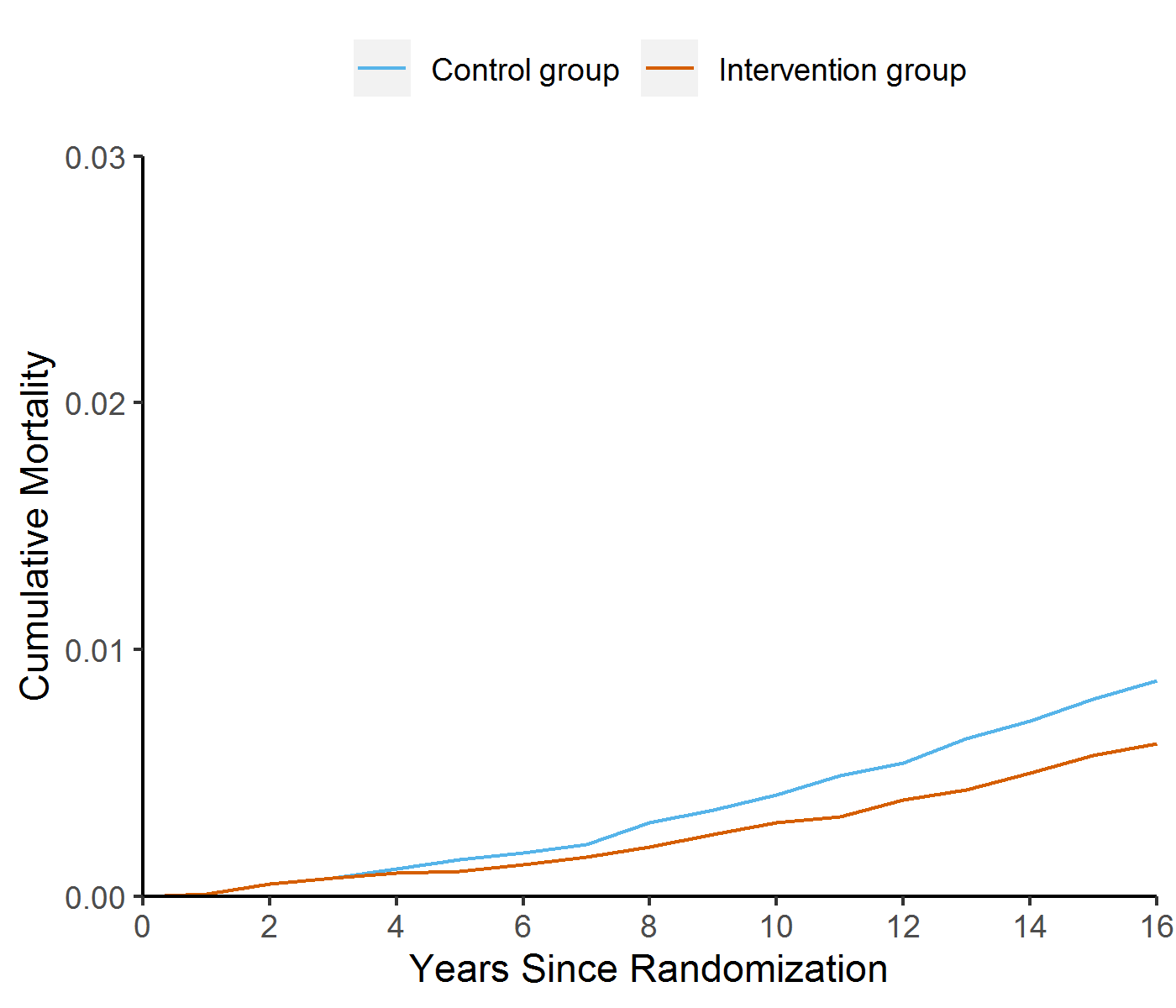 |
| **PLCO^4^** | |
| 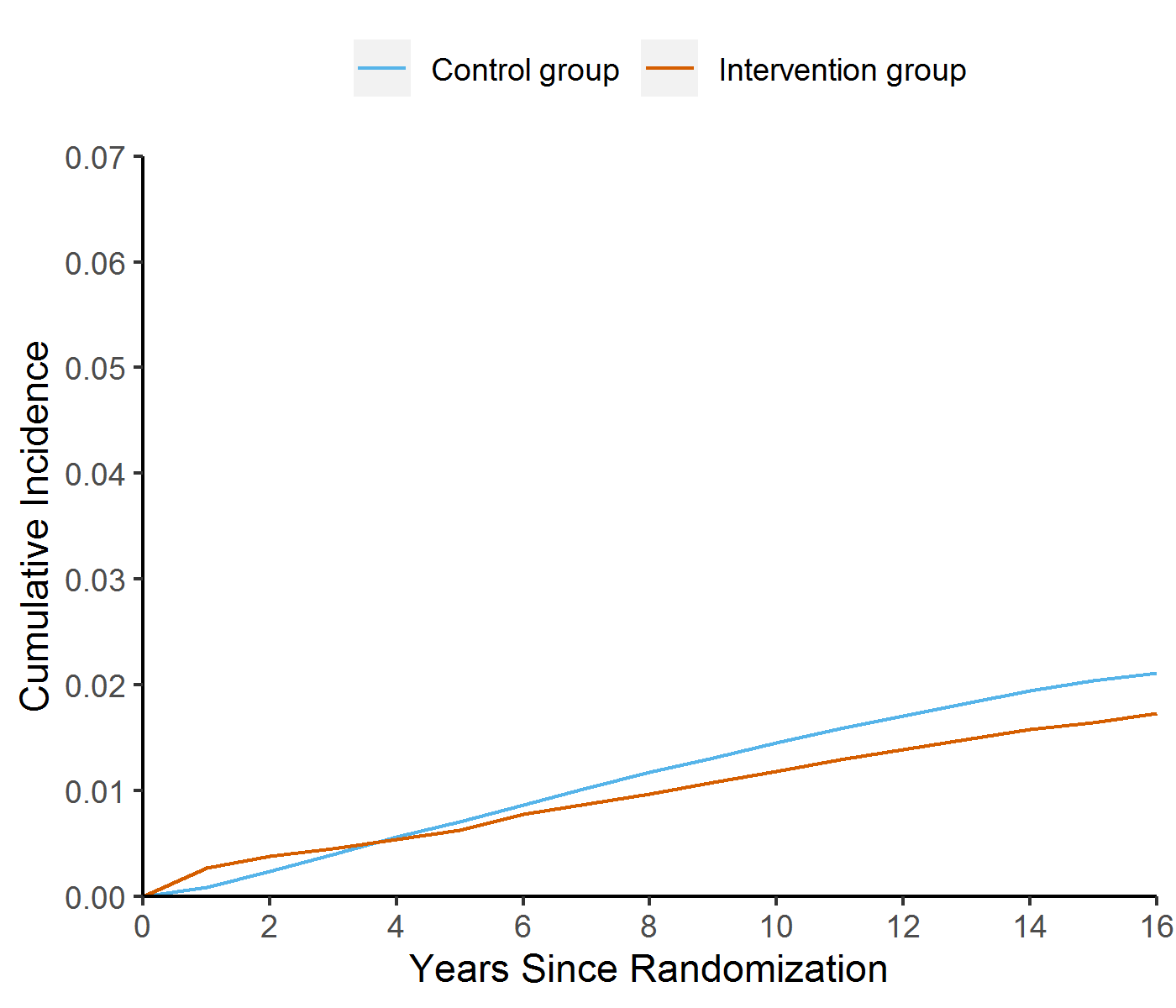 | 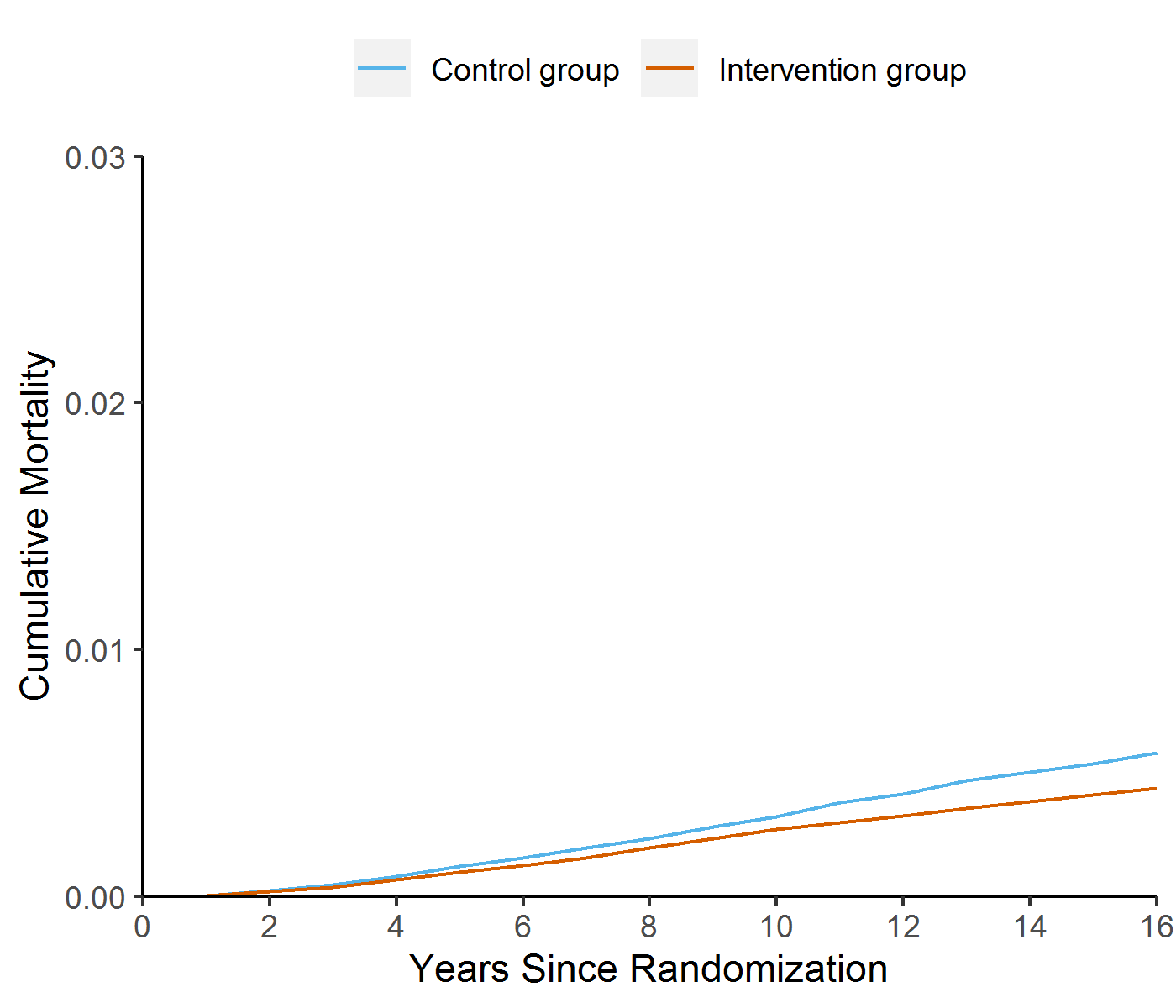 |
| ^1^ approximated using data extracted from reference ^3^  ^2^ approximated using data extracted from reference ^4^  ^3^ approximated using data extracted from reference ^5^  ^4^ approximated using data extracted from reference ^6^ | |

#### **Supplementary Figure 4.** Annual incidence (top) and mortality (bottom) rate ratios (screening /control).

**Per Protocol**

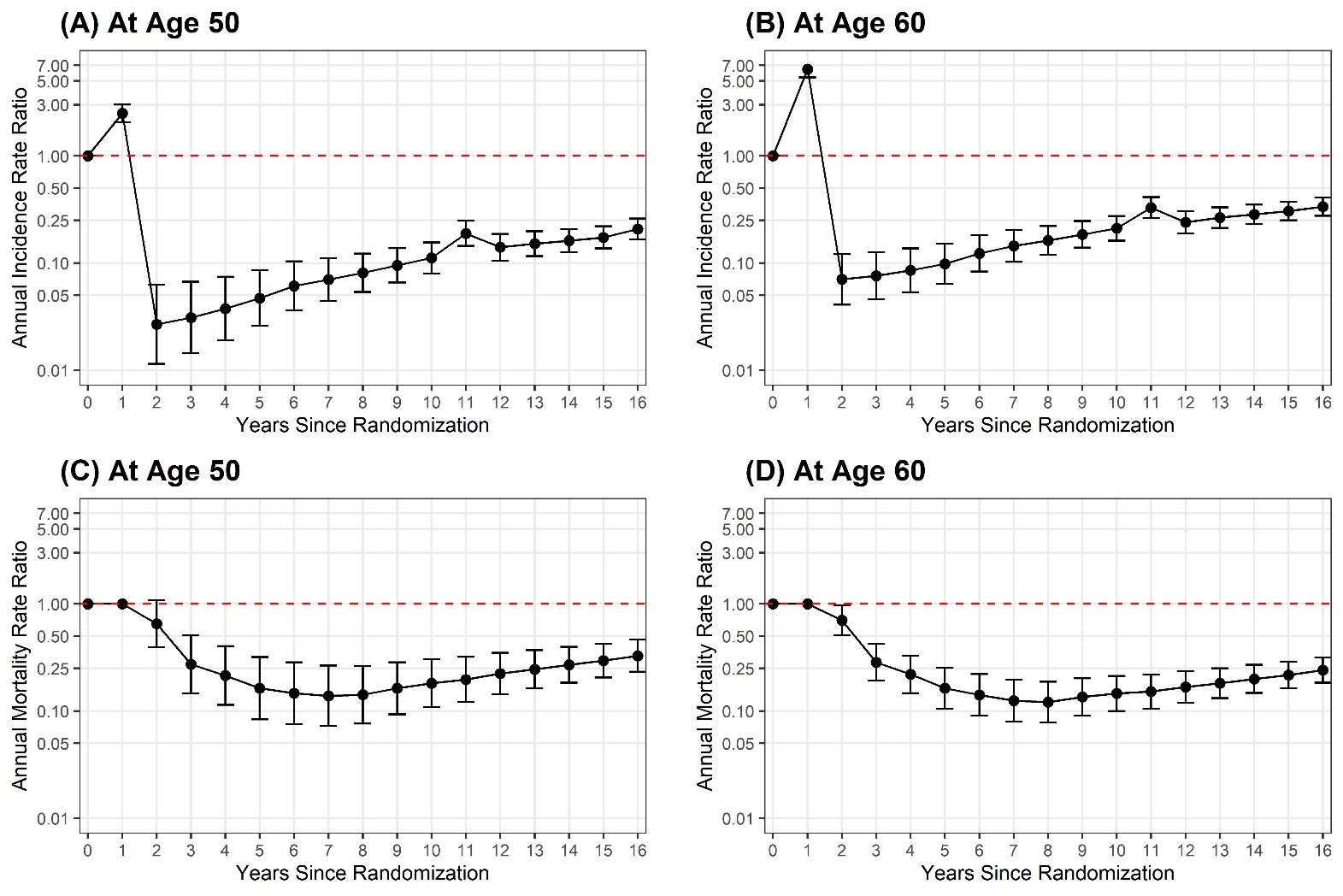

**Intention to Screen**

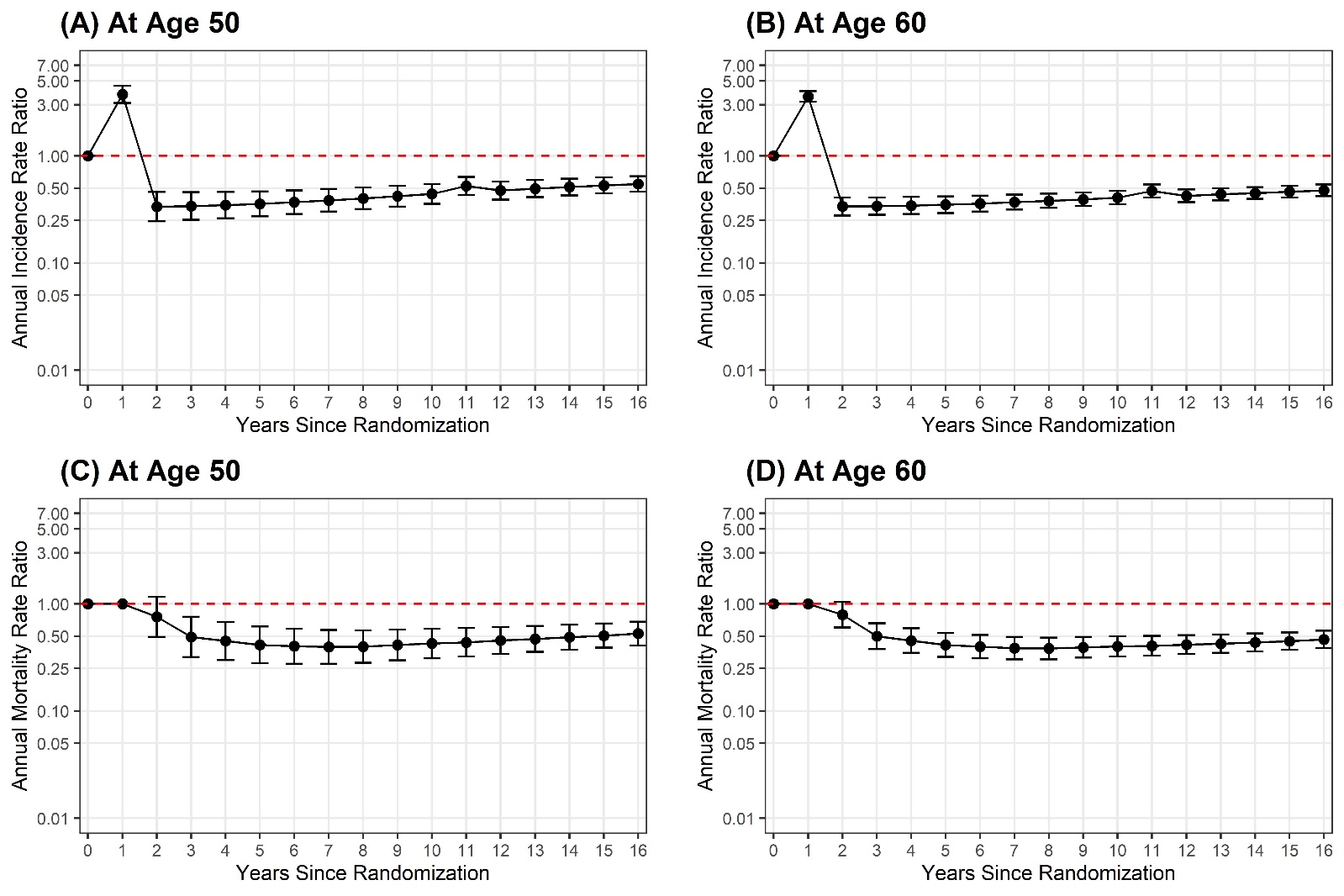

#### **Supplementary Figure 5.** Cumulative incidence at different starting ages of the simulation, stratified by sex.

**Per Protocol**

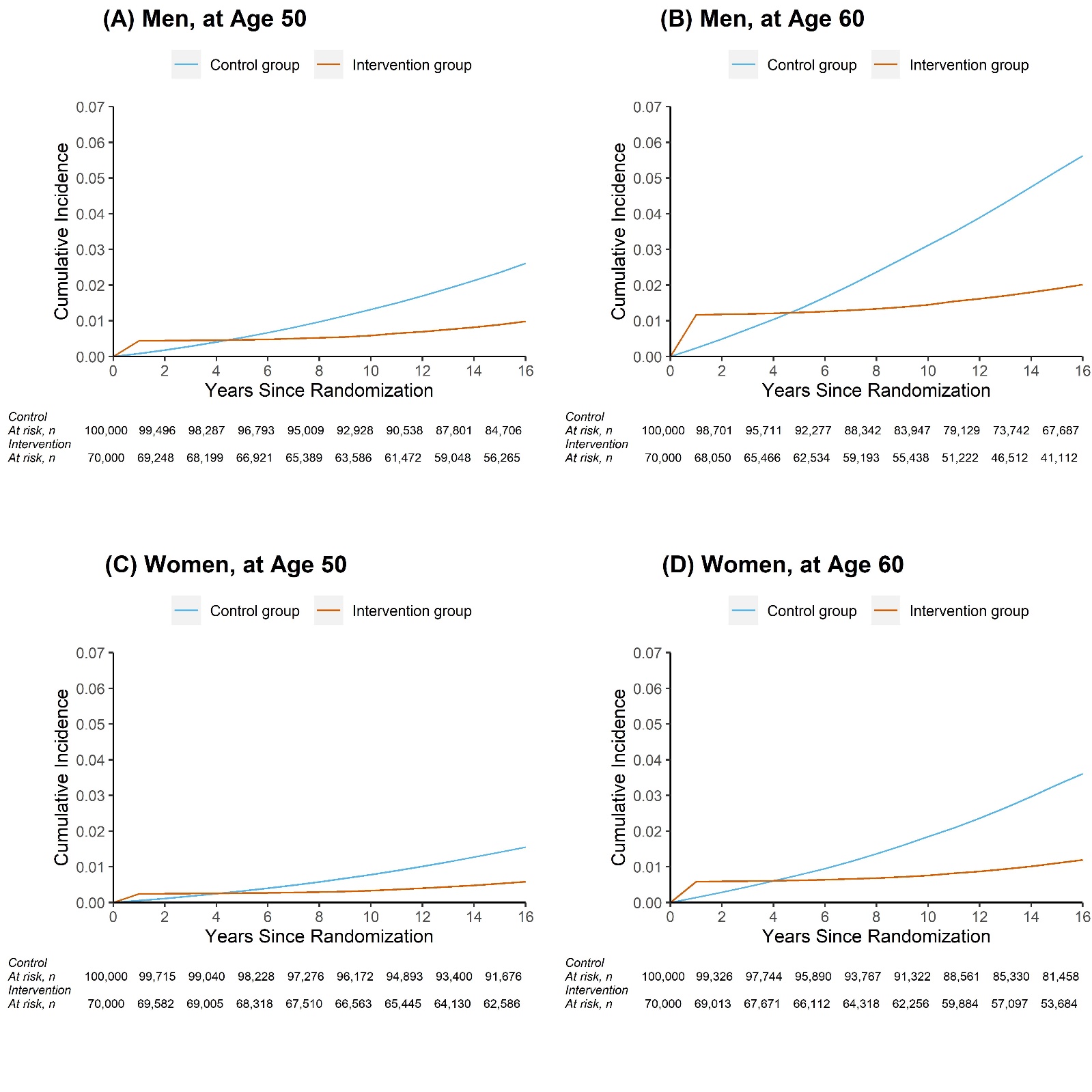

**Intention to Screen**

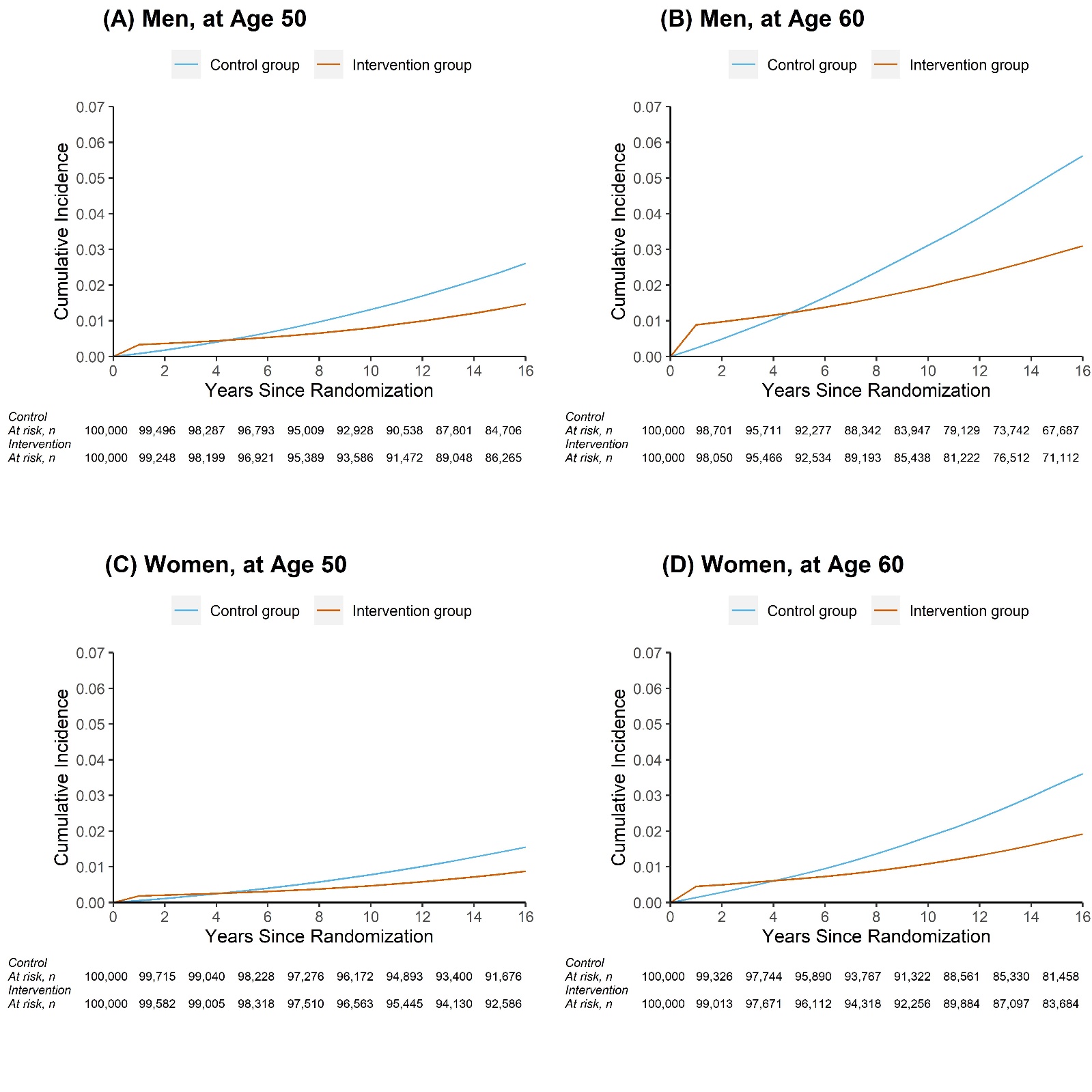

#### **Supplementary Figure 6.** Cumulative mortality at different starting ages of the simulation, stratified by sex.

**Per Protocol**

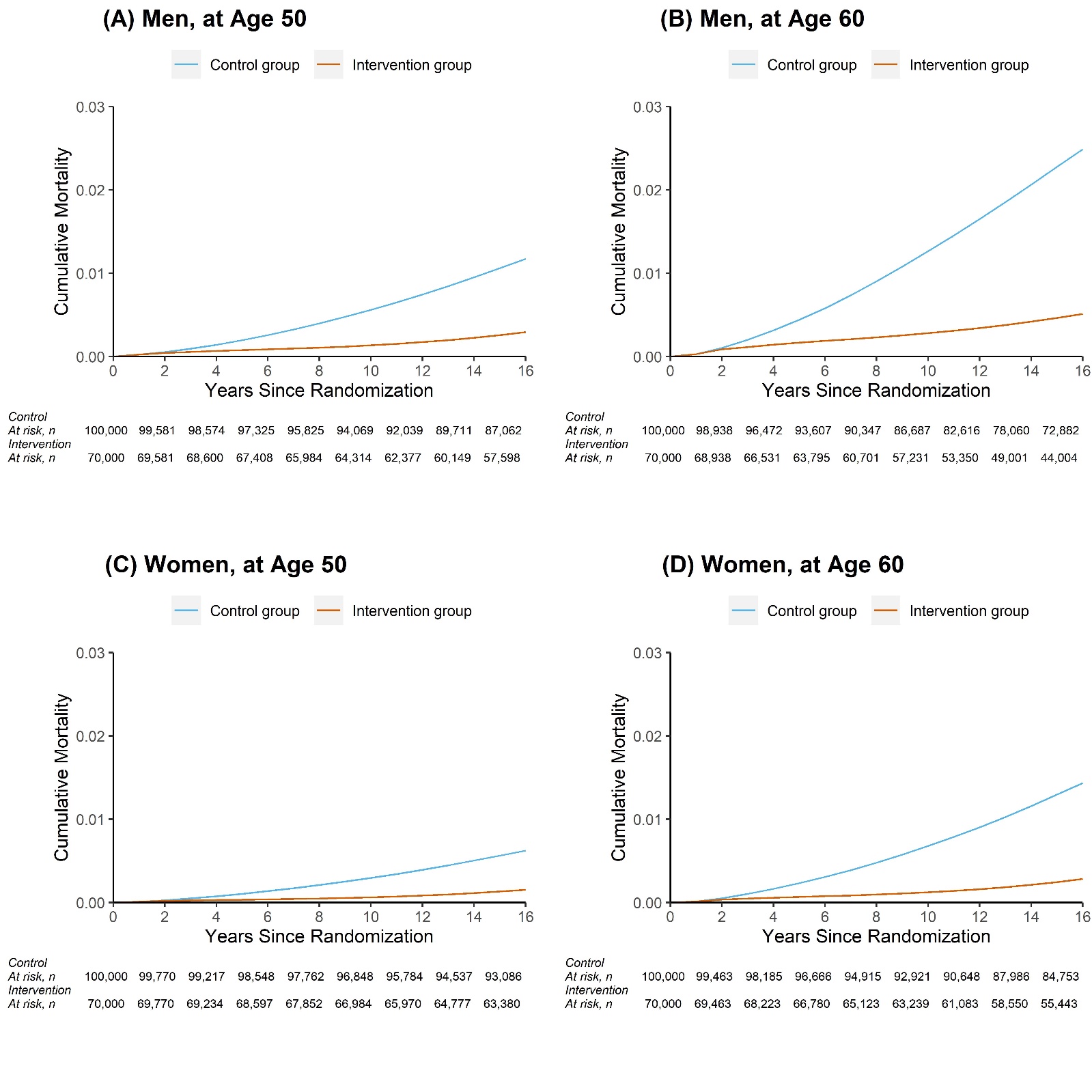

**Intention to Screen**

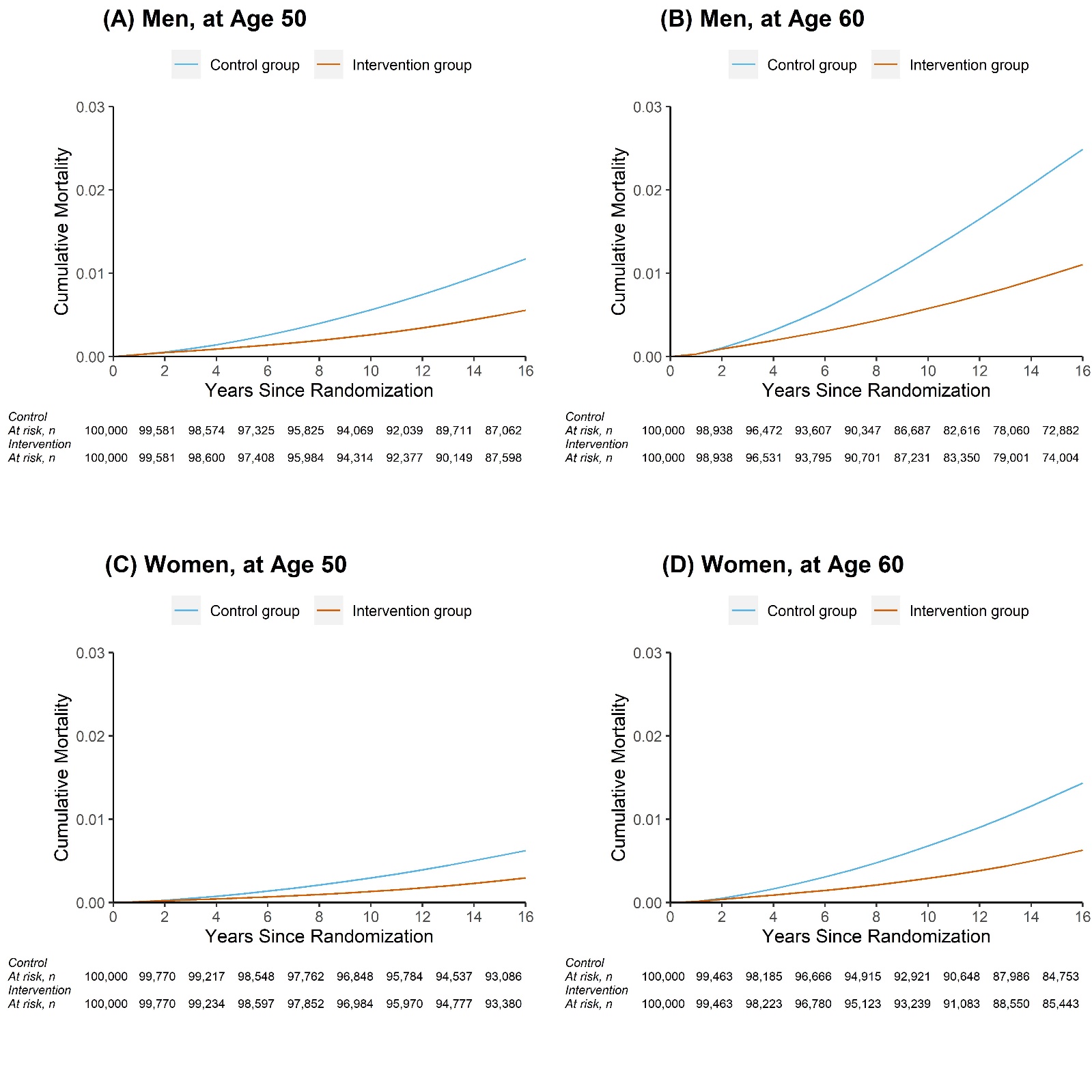

#### **Supplementary Figure 7.** Annual incidence rate ratios (screening /control), stratified by sex.

**Per Protocol**

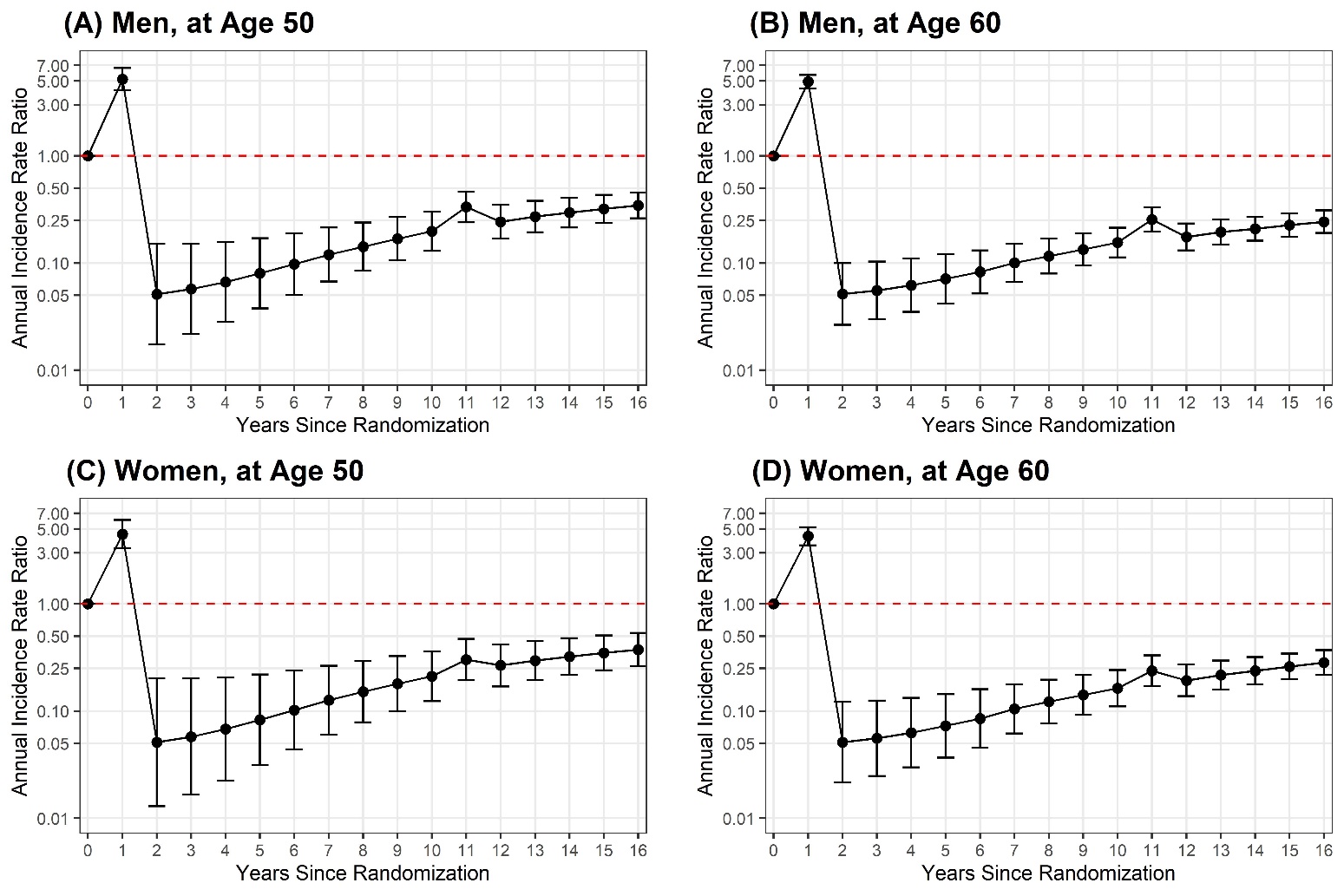

**Intention to Screen**

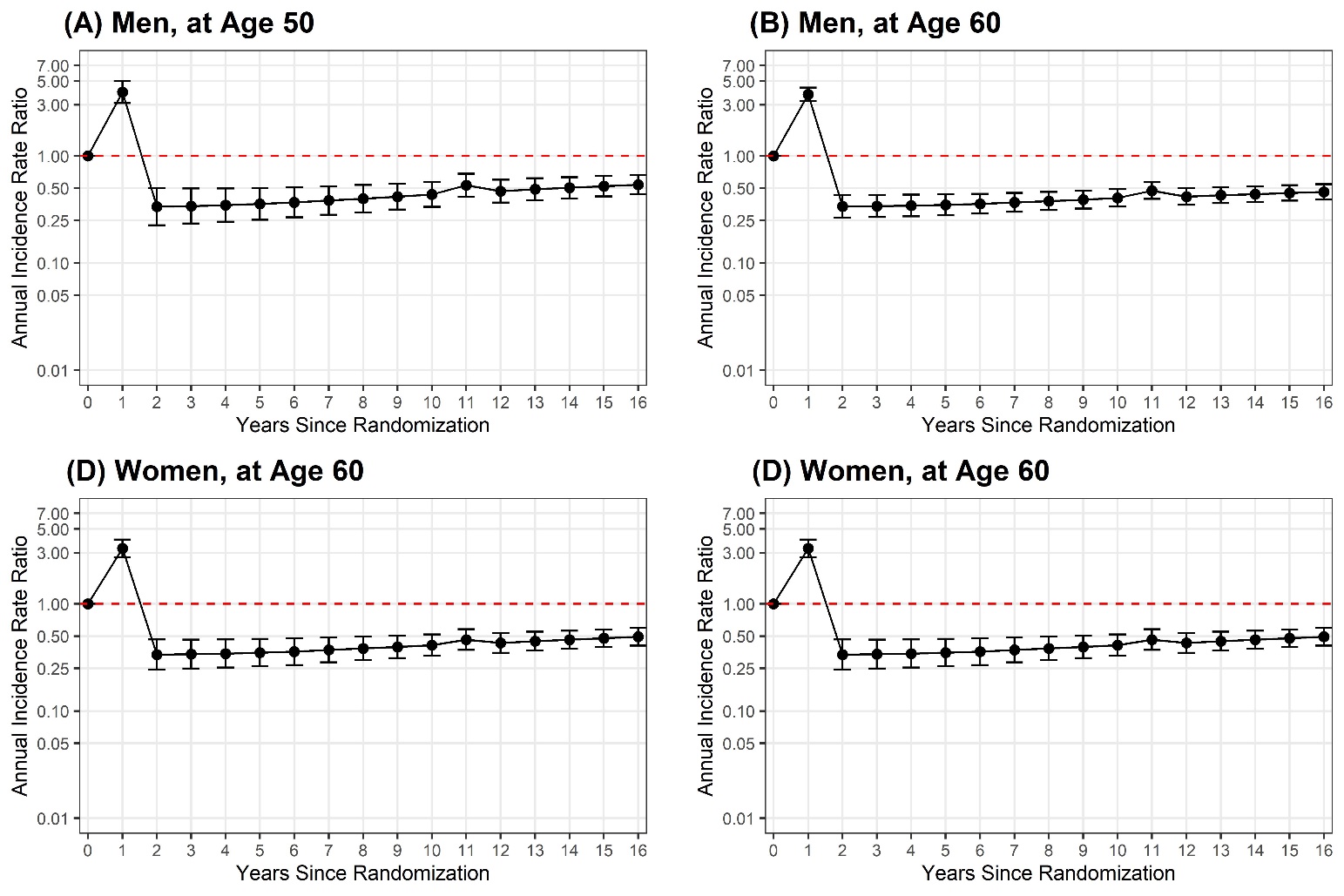

#### **Supplementary Figure 8.** Annual mortality rate ratios (screening /control), stratified by sex.

**Per Protocol**

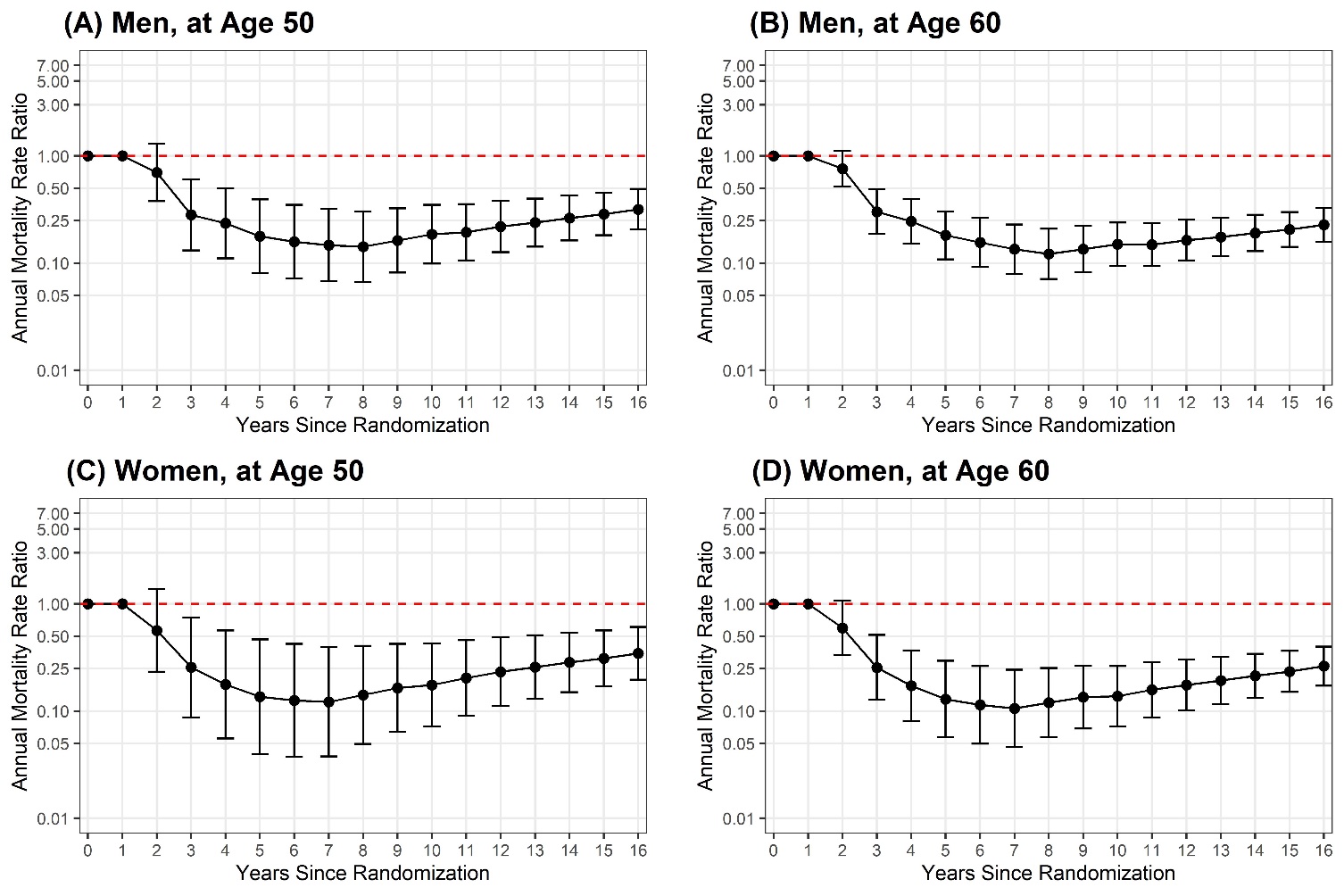

**Intention to Screen**

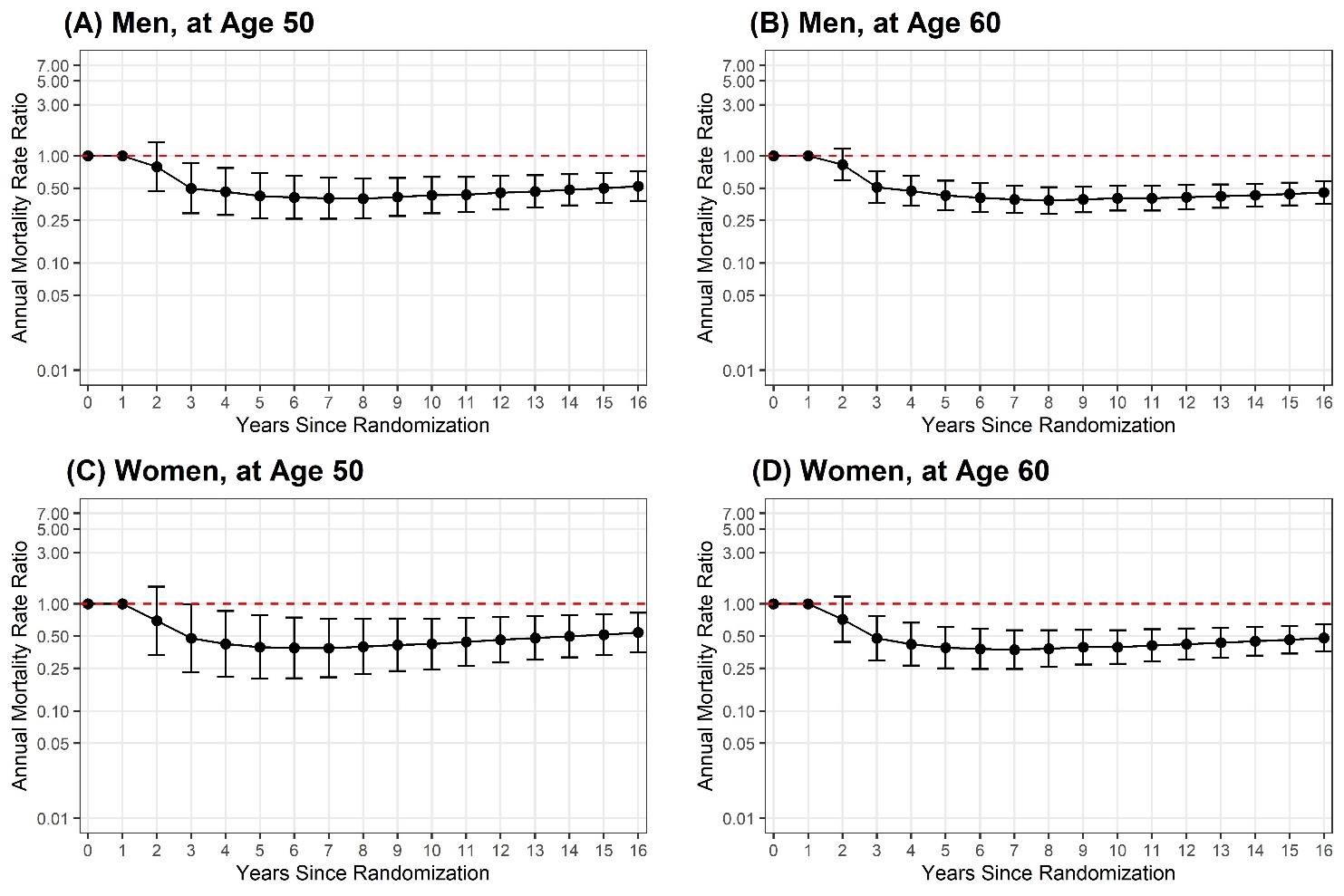
